# Heterogeneous Effect of Automated Alerts on Mortality

**DOI:** 10.1101/2025.08.11.25333302

**Authors:** Benjamin D. Wissel, Zana Percy, Tanner J. Zachem, Brett Beaulieu-Jones, Isaac S. Kohane, Stuart L. Goldstein, Emrah Gecili, Judith W. Dexheimer

**Affiliations:** Division of Biomedical Informatics, Cincinnati Children’s Hospital Medical Center, Cincinnati, OH, USA; Department of Neurosurgery, Duke University, Durham, NC, USA; Department of Environmental and Public Health Sciences, University of Cincinnati College of Medicine, Cincinnati, OH, USA; Department of Biomedical Informatics, Harvard Medical School, Boston, MA, USA; Section of Biomedical Data Science, Department of Medicine, University of Chicago, Chicago, IL, USA; Department of Pediatrics, University of Cincinnati College of Medicine, Cincinnati, OH, USA; Center for Acute Care Nephrology, Cincinnati Children’s Hospital Medical Center, Cincinnati, OH, USA; Division of Biostatistics and Epidemiology, Cincinnati Children’s Hospital Medical Center, Cincinnati, OH, USA; Division of Emergency Medicine, Cincinnati Children’s Hospital Medical Center, Cincinnati, OH, USA

**Keywords:** clinical decision support, meta-analysis, medical informatics, acute kidney injury

## Abstract

We analyzed data from 13,483 hospitalized patients with acute kidney injury (AKI) from three randomized controlled trials to assess the heterogeneous effects of automated electronic alerts on 14-day mortality. We modeled and predicted individualized alert effects on a subset of the ELAIA-1 patients and validated it internally on ELAIA-1 holdout patients and externally on ELAIA-2 and UPenn trial patients. Patients predicted to benefit from alerts had significantly lower mortality compared to those predicted to be harmed (p-interaction<0.05). In external cohorts, 43 deaths may have been preventable if alerts were restricted to likely beneficiaries. Machine-learning based meta-analysis identified reduced mortality with alerts among patients with higher blood pressures and lower predicted risk, but increased mortality in non-urban and non-teaching hospitals. Provider responses to alerts varied across subgroups. These findings suggest that tailoring alerts to patient phenotypes may improve outcomes and support the need for a prospective trial of individualized alert strategies.

**Trial Registration:** https://clinicaltrials.gov/ct2/show/NCT02753751 and https://clinicaltrials.gov/ct2/show/NCT02771977

## INTRODUCTION

Acute kidney injury (AKI) is a syndrome that results in a sudden decline in kidney function.^1^ It occurs in 21-26% of hospitalized patients and carries a mortality rate of 21-24%.^2,3^ A substantial portion of these deaths could be prevented with improved clinical care.^4^ A review by the National Confidential Enquiry into Patient Outcome and Death found that only 50% of hospitalized patients with AKI received care that adhered to established guidelines. There were unacceptable delays in recognizing 43% of AKIs that developed during hospital admissions.^4^

Automated alerting systems aim to improve the identification of AKI, increase adherence to treatment guidelines, and reduce the delay to treatment. Providers who receive AKI alerts are more likely to order intravenous (IV) fluids, a urinalysis, measure serum creatinine, and document AKI in the problem list.^5^ Large healthcare systems, including the National Health System in England, have implemented automated alerting systems for in-hospital AKI.^6^ However, clinical studies on alert systems have yielded mixed results on patient outcomes.^6,7^ One large, double-blind, multicenter, parallel-group, randomized controlled trial found that electronic health record-based (EHR) alerts had heterogeneous treatment effects across hospitals.^5^ Patients at non-teaching hospitals who were randomized to the alert group had an 81% higher risk of death than those in the control group, corresponding to 26 excess deaths in the trial. Conversely, there was a non-significant trend toward a reduced risk of death after AKI alerts at teaching hospitals.

We hypothesized that patient characteristics and hospital settings modify the effect of alerts on mortality. To investigate, we used machine learning to predict the individualized treatment effects of alerts using data from two randomized controlled trials.^5,8^ First, we mapped patients from the <u>El</u>ectronic <u>A</u>lerts for <u>A</u>cute Kidney Injury <u>A</u>melioration (ELAIA-1) trial to phenotypic signatures based on their baseline clinical characteristics. We leveraged information from patients’ intervention allocation (alert vs. usual care) and their subsequent outcomes to predict the effect that randomization to the alert group had on their risk of mortality, using a holdout dataset for internal validation.^5^ Second, we externally validated the model on patients in two additional randomized controlled trials: one conducted at the University of Pennsylvania (UPenn trial)^8^ and the Electronic Alerts for AKI Amelioration 2 (ELAIA-2) trial.^9^ Third, we pooled individual patient data from the three trials for a meta-analysis that tested which clinical characteristics were associated with a higher risk of mortality following randomization to the alert group. Finally, we analyzed provider actions as a possible mechanism for heterogeneous treatment effects following automated alerts.

## RESULTS

### Description of the study population

Data from all 6,030 patients in the ELAIA-1 trial (n=4,221 in the training cohort, and n=1,809 in the interval validation cohort), 5,060 patients in the ELAIA-2 trial (external validation cohort), and 2,393 patients in the UPenn trial (external validation cohort) were used in this intention-to-treat analysis. A total of 36 studies were screened for eligibility, six studies underwent full text screening, and three were included in the final study (Supplemental Figure 1 Supplementary Note 1). The study was conducted in accordance with the Preferred Reporting Items for Systematic reviews and Meta-Analyses extension for Individual Participant Data (PRISMA-IPD, Supplementary Note 2). All three trials were high-quality and judged to have a low risk of bias according to the Cochrane Risk of Bias.^10^ Patient characteristics varied across trials (Table 1). Compared to patients in the UPenn trial, patients in the ELAIA-1 trial were older and more likely to have diabetes, chronic heart failure, higher creatinine, and lower blood pressure (all p<0.001). Mortality was higher in the ELAIA trials than the UPenn trial (8.9% and 11% vs. 7.1%, p<0.001). Factors contributing to patient mortality were heterogeneous across patients and hospitals (Supplementary Figure 2). Visualizing patients by the factors that contributed to their predicted risk of death revealed distinctive patterns (Figure 1 and Supplementary Figures 3 and 4). Patients in hospitals 4 and 5, patients in the ICU, patients exposed to nephrotoxic drugs (NSAIDs, RAS-inhibiting agents, and aminoglycosides), and patients with chronic kidney disease clustered together. These clusters revealed subgroups of patients whose risk of death was attributable to similar factors. There were 2,161 (0.3%) missing data points in the ELAIA-1 trial, 48 (0.04%) missing data points in the ELAIA-2 trial, and 3,692 (11.0%) missing data points in the UPenn trial. In the ELAIA-1 trial, 23 (0.4%) patients had missing blood pressure values, and two (0.03%) patients had missing creatinine values. In the ELAIA-2 trial, 12 (0.2%) patients were missing blood pressure values, and 13 (0.03%) patients were missing follow up time. In the UPenn trial, 1,826 (76%) patients had missing blood pressure values, 17 (0.7%) patients were missing age and gender, and two (0.1%) patients were missing chronic heart failure and diabetes comorbidity statuses.

**Table 1.** Baseline trial participant characteristics. Data from all 13,483 patients were reported for all characteristics. Patient race was not included as part of the ELAIA-2 trial. N=number of patients; ELAIA-1=Electronic Alerts for Acute Kidney Injury Amelioration-1 trial; ELAIA-2=Electronic Alerts for Acute Kidney Injury Amelioration-2 trial; and UPenn=University of Pennsylvania trial.

| Characteristic | Overall,<br>N = 13,483<sup>1</sup> | ELAIA-1,<br>N = 6,030<sup>1</sup> | ELAIA-2,<br>N = 5,060<sup>1</sup> | UPenn,<br>N = 2,393<sup>1</sup> |
| --- | --- | --- | --- | --- |
| **Group** |  |  |  |  |
| Alert | 6,792 (50%) | 3,059 (51%) | 2,532 (50%) | 1,201 (50%) |
| Control | 6,691 (50%) | 2,971 (49%) | 2,528 (50%) | 1,192 (50%) |
| Deaths within 14 days | 1,250 (9.3%) | 537 (8.9%) | 535 (11%) | 178 (7.4%) |
| **Demographics** |  |  |  |  |
| Age (Years) | 69 (58, 80) | 71 (59, 82) | 70 (59, 80) | 62 (52, 72) |
| Sex |  |  |  |  |
| Female | 6,392 (47%) | 2,882 (48%) | 2,453 (48%) | 1,057 (45%) |
| Male | 7,091 (53%) | 3,148 (52%) | 2,607 (52%) | 1,336 (55%) |
| Black or African American | 1,593 (19%) | 946 (16%) | - | 647 (27%) |
| **Hospital Type** |  |  |  |  |
| Teaching Hospital | 12,718 (94%) | 5,265 (87%) | 5,060 (100%) | 2,393 (100%) |
| Urban Hospital | 11,662 (86%) | 4,698 (78%) | 4,571 (90%) | 2,393 (100%) |
| ICU | 3,803 (28%) | 1,923 (32%) | 1,158 (23%) | 722 (30%) |
| **Previous Diagnoses** |  |  |  |  |
| Chronic Heart | 5,029 (37%) | 2,658 (44%) | 1,611 (32%) | 760 (32%) |
| Diabetes | 5,102 (38%) | 2,484 (41%) | 1,895 (37%) | 723 (30%) |
| Malignancy | 2,728 (20%) | 931 (15%) | 1,112 (22%) | 685 (29%) |
| **Lab Values** |  |  |  |  |
| Systolic Blood Pressure (mmHg) | 121 (107, 136) | 118 (104, 134) | 118 (104, 134) | 131 (124, 139) |
| Diastolic Blood Pressure (mmHg) | 69 (60, 77) | 66 (58, 75) | 67 (59, 76) | 75 (70, 80) |
| Baseline Creatinine (mg/dL) | 0.98 (0.70, 1.40) | 1.01 (0.71, 1.44) | 0.97 (0.70, 1.39) | 0.90 (0.59, 1.37) |
| Creatinine at Randomization (mg/dL) | 1.48 (1.12, 1.98) | 1.50 (1.15, 2.00) | 1.48 (1.15, 1.98) | 1.37 (0.98, 1.90) |
| Change in Creatinine (mg/dL) | 0.40 (0.32, 0.54) | 0.40 (0.32, 0.56) | 0.40 (0.31, 0.55) | 0.37 (0.31, 0.50) |

<sup>1</sup>n (%); Median (IQR).

**Figure 1.**
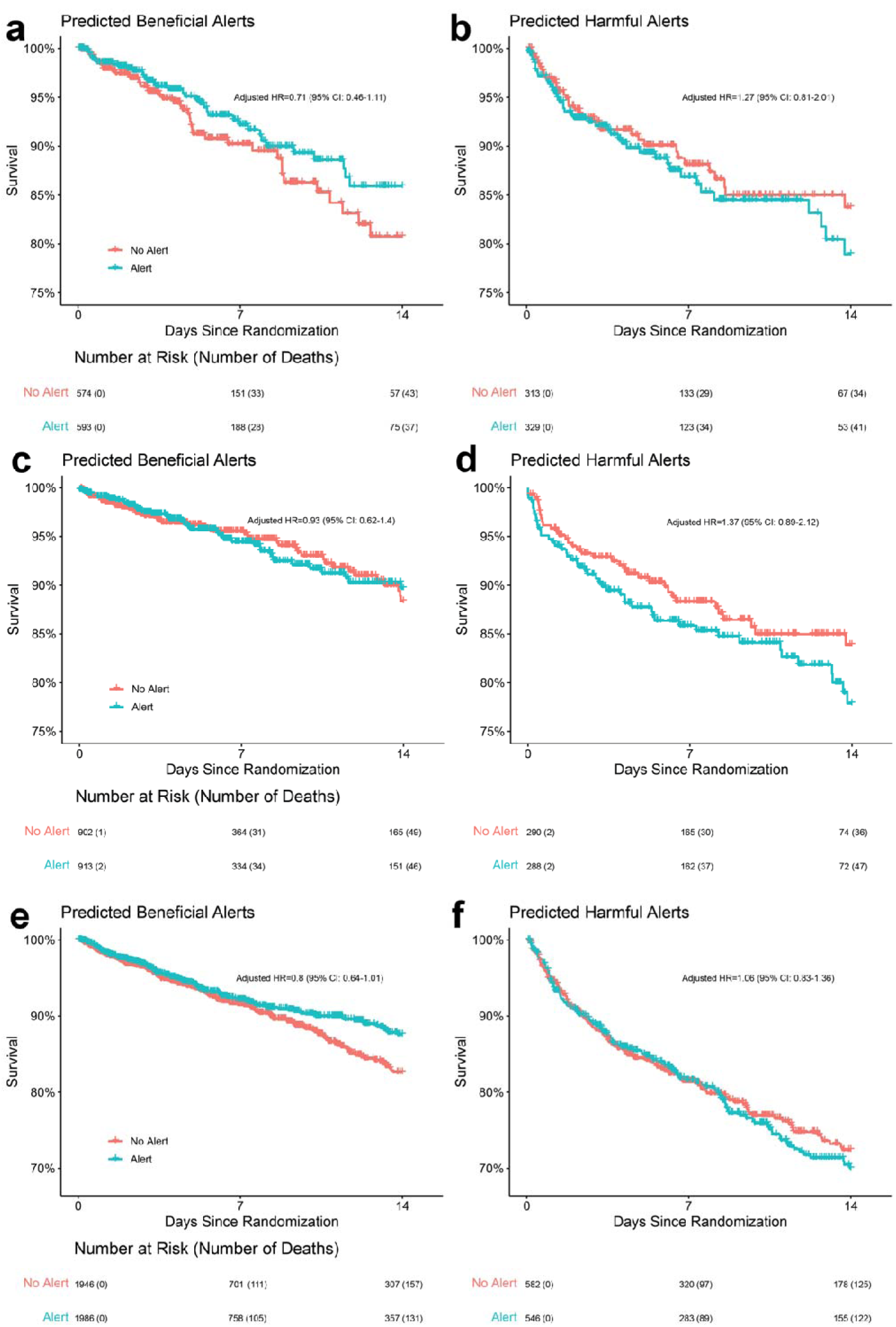
Performance of the model on a holdout dataset from the ELAIA-1 trial (A and B, top; internal validation), UPenn trial (C and D, middle; external validation), and ELAIA-2 trial (E and F, bottom; external validation). Hazard ratios (iHR) were adjusted for patients’ baseline risk of death.

### Predicting individualized risk of death following an automated alert

Among the 30% of patients in the ELAIA-1 trial who were in the holdout dataset, the median individualized hazard ratio (iHR) for death following an alert was 0.84 (IQR: 0.69-1.14), indicating that alerts were predicted to be beneficial in most patients. There were 1,167 (64.5%) patients predicted to benefit from an alert (iHR<1) and 642 (35.5%) patients predicted to be harmed (iHR>1). In our external validation, the median iHR for death following an alert among patients in the UPenn trial was 0.73 (IQR: 0.61-0.98). Among the 2,393 UPenn trial patients, 1,815 (75.8%) were predicted to benefit from an alert, while 578 (24.2%) were predicted to be harmed. The median iHR for death following an alert among patients in the ELAIA-2 trial was 0.77 (IQR: 0.64-0.95). Among the 5,060 ELAIA-2 trial patients, 3,932 (77.7%) were predicted to benefit from an alert, while 1,128 (22.3%) were predicted to be harmed.

Within the group of patients that were randomized to receive alerts in the ELAIA-1 trial (internal validation cohort), those predicted to benefit from an alert had a lower risk of death than patients who were predicted to be harmed by an alert (adjusted HR=0.71 [95% CI: 0.46−1.11] vs. adjusted HR=1.27 [95% CI: 0.81−2.01]; p_interaction_=0.045) (Figure 1). In the UPenn trial (external validation), patients predicted to benefit from an alert had a lower risk of death than patients who were predicted to be harmed by an alert (HR=0.93 [95% CI: 0.62−1.40] vs. HR=1.37 [95% CI: 0.89−2.12]; p_interaction_=0.009). In the ELAIA-2 trial (external validation), patients predicted to benefit from an alert had a lower risk of death than patients who were predicted to be harmed by an alert (HR=0.80 [95% CI: 0.64−1.01] vs. HR=1.06 [95% CI: 0.83−1.36]; p_interaction_<0.0001).

Among patients in the external validation cohorts who were predicted to benefit from an alert, there were 29 fewer deaths in patients who were randomized to the alert group. Among patients predicted to be harmed by an alert, there were 14 excess deaths in patients who were randomized to the alert group (Figure 1). In total, there would have been 43 fewer deaths in the two external validation trials (n=7,453), corresponding to a 6.0% (95% CI: 5.8-6.3%) reduction in the relative risk of all-cause mortality, and 0.58% (95% CI: 0.56-0.60%) reduction in the absolute risk of all-cause mortality. iHRs>3 were clipped at 3 (n=8 [0.1%] patients) and iHRs<0.33 (n=104 [1.4%] patients) were clipped at 0.33, based on the assumption that alerts could not impact patient mortality by greater than 3-fold. We performed a sensitivity analysis that did not clip iHRs, and the results did not change.

The top 5 most important features that impacted patients’ iHRs were ICU status, hospital teaching status, CHF, diabetes, and sex (Figure 2). Patients in the ICU were predicted to have a higher risk of mortality following randomization to the alert group, though the magnitude of this interaction depended upon their phenotypic signature (Supplementary Figure 5). Our model captured non-linear relationships between predicted iHR and patient age, blood pressure, creatinine, CHF comorbidity, and race (Figure 2), highlighting the heterogeneity of individualized treatment effects across phenotypic signatures. We released the trained XGBoost model on GitHub (https://github.com/wisselbd/AKI_Alerts).

**Figure 2.**
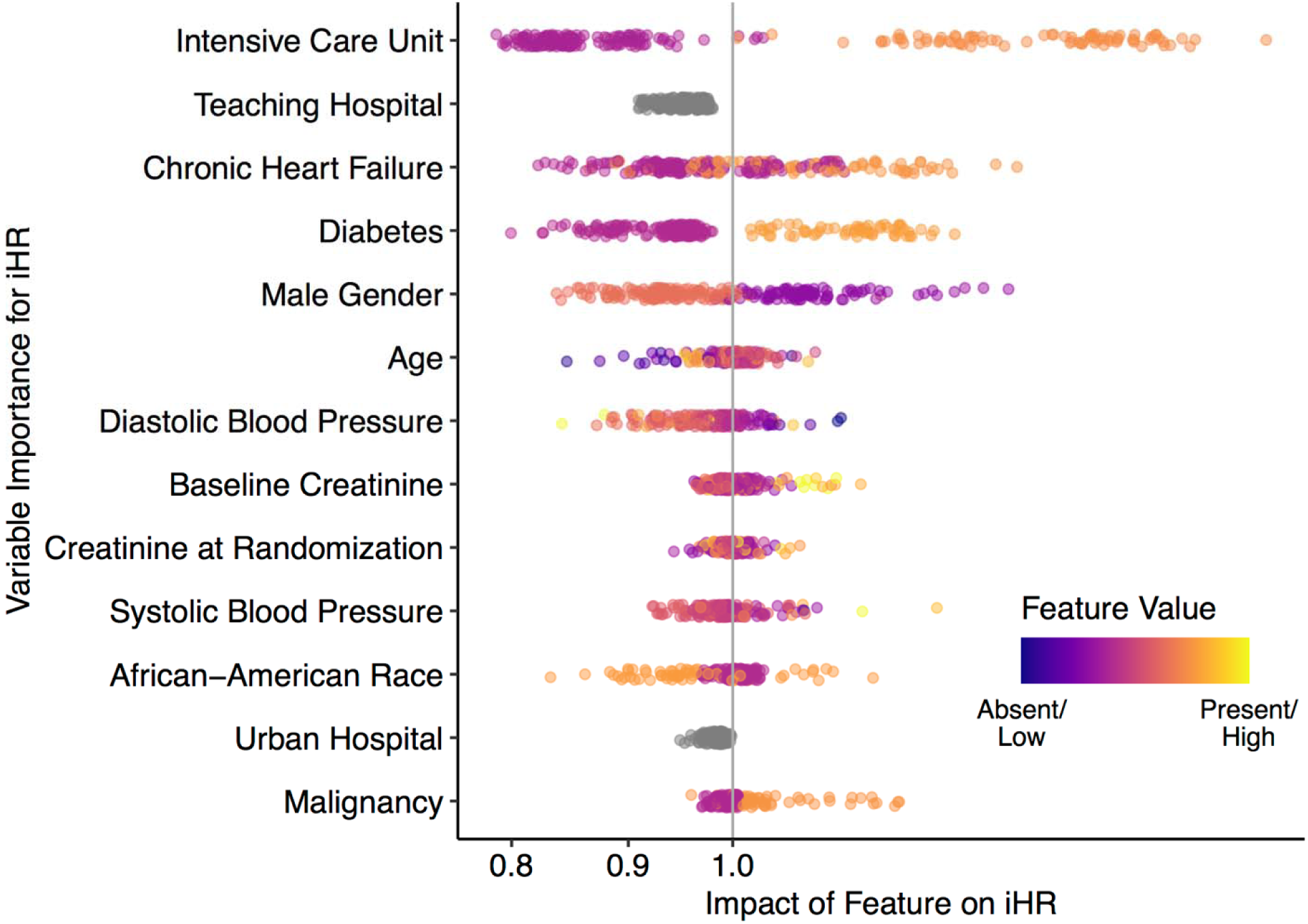
Summary of variable impact on patients’ predicted benefit from an alert. Variables at the top (e.g. intensive care unit) had the biggest impact on the alerts predicted effect on mortality. Variables at the bottom (e.g. malignancy) had the smallest impact. Each dot represents one patient. Patients with low feature values were colored in purple, while high feature values were colored in yellow. SHAP values (x-axis, log scale) correspond to the features’ impact on the predicted effect of alerts. Variables at the top had the largest impact, on average, while variables at the bottom had the lowest impact on the predicted effect of alerts. iHR: individualized hazard ratio.

### Association between alerts and mortality across patient subgroups

In the meta-analysis of individual patient data from the ELAIA-1, ELAIA-2, and UPenn trials, the τ^2^ value was estimated at 0.0002 (95% CI: 0.00-0.0202), with an I^2^ value of 2.20%, indicating very low between-study heterogeneity in the distribution and variability of true effect sizes (p=0.34). At the population level, alerts were not associated with the risk of all-cause mortality (n=3 studies, n=13,483 patients, 618 deaths [9.10%] in the alert group vs. 632 deaths [9.45%] in the control group, adjusted HR=0.96 [95% CI: 0.86-1.07], p=0.47) (Table 1). Patient characteristics were balanced between alert and control groups for the entire population (Supplementary Table 1) and for patient subgroups (Supplementary Tables 2-4).

Alerts were associated with a decreased risk of mortality in patients with higher systolic blood pressures at baseline (SBP >130; n = 4,551 patients, 4.5% vs 5.3% deaths, HR = 0.83 [95% CI: 0.66-1.04]). In contrast, alerts were associated with an increased risk of mortality in patients with low systolic blood pressures (SBP ≤ 90; n = 683 patients, 27.5% vs 24.0% deaths, HR = 1.43 [95% CI: 1.11-1.83]), in suburban hospitals (n = 1,821 patients, 14.2% vs 10.9% deaths, HR = 1.34 [95% CI: 1.03-1.74]), and in non-teaching hospitals (n = 765 patients, 15.6% vs 8.5% deaths, HR = 1.96 [95% CI: 1.28-3.00]). Additionally, alerts were favored in patients whose predicted risk of death was in the lowest quartile (n = 3,371, 22.4% vs 21.0% deaths, HR = 0.61 [95% CI: 0.41-0.91]; Figure 3).

**Figure 3.**
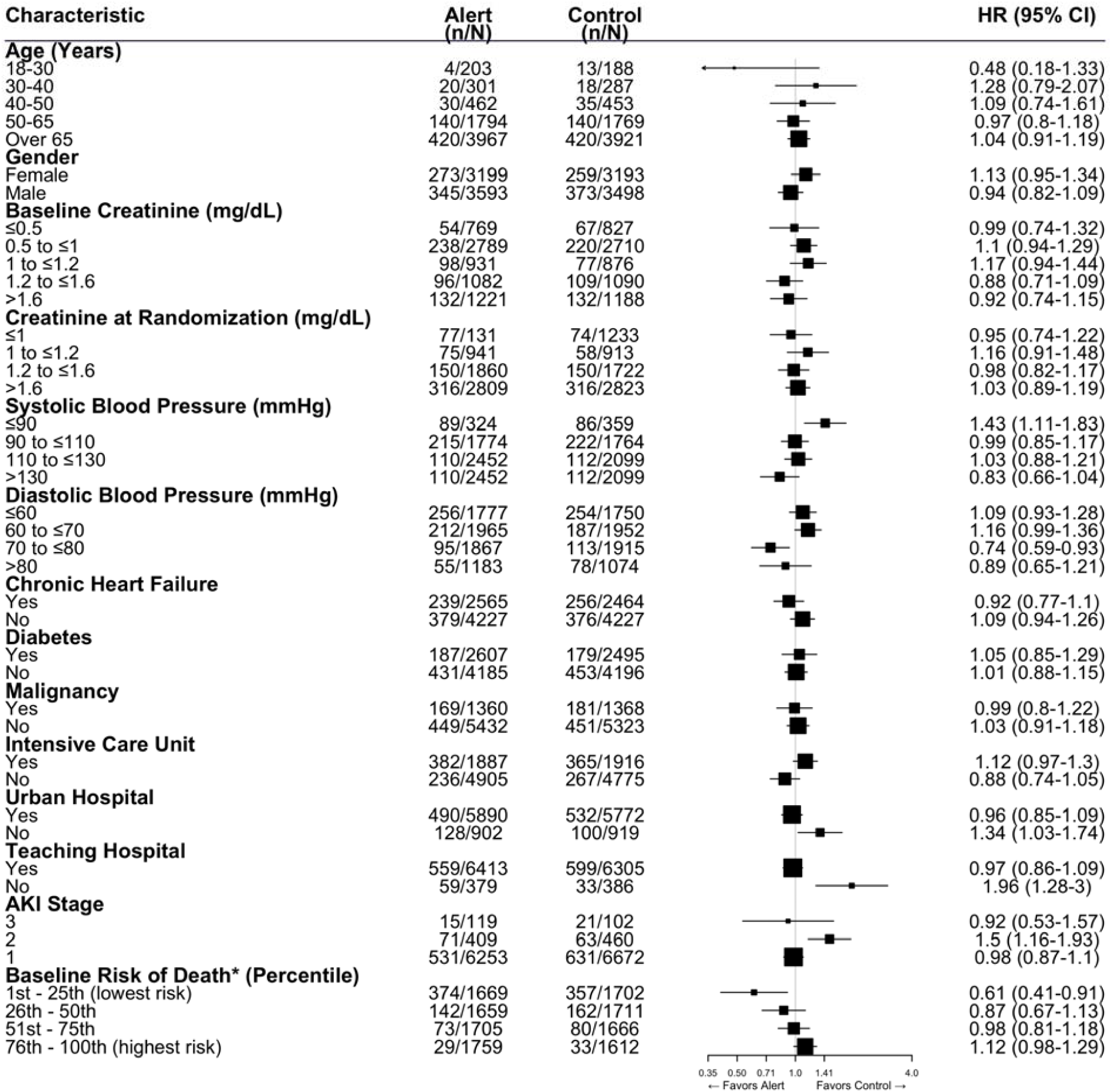
Association between alerts and all-cause mortality in patient subgroups using individual patient data from the ELAIA-1, ELAIA-2, and UPenn trials. *Baseline risk of death includes data from UPenn and ELAIA-2 trials only. n=number of deaths; N=number of patients; CI=confidence interval; and HR=hazard ratio.

Age, gender, creatinine at randomization, prior diagnosis of diabetes, heart failure, history of malignancy, and ICU status were not associated with the effect of alerts on mortality (Figure 3). We observed batch effects across two of the seven individual hospitals (p<0.05 the interaction between alert and two of the individual hospitals). Alerts were associated with a decreased risk of mortality at two urban, teaching hospitals (Hospital 1: adjusted HR=0.68 [95% CI: 0.47-0.96], p=0.031; and Hospital 3: adjusted HR=0.54 [95% CI: 0.35-0.82], p=0.0039].

Patients predicted to benefit from alerts had a lower baseline risk of death than patients who were not predicted to benefit (median=0.05 [IQR: 0.03-0.09] vs. median=0.12 [IQR: 0.07-0.19]; unadjusted p<0.001) (Table 2). However, after adjusting for other factors, the association between baseline risk of death and the effect of alerts was not significant (adjusted HR=1.01 per 10% absolute risk of death [95% CI: 0.75-1.36], p=0.95) (Table 2). Similarly, AKI stage did not mediate the effect of alerts on mortality (adjusted HR=0.79 [95% CI: 0.56-1.13], p=0.21).

**Table 2.** Characteristics of patients predicted to benefit from an alert in the two external validation studies.

| Characteristic | N | Alert Beneficial, N<br>= 5,747<sup>1</sup> | Alert Not Beneficial, N<br>= 1,706<sup>1</sup> | p-value<sup>2</sup> |
| --- | --- | --- | --- | --- |
| **Study** | 7,453 |  |  | 0.074 |
| ELAIA-2 |  | 3,932 (68%) | 1,128 (66%) |  |
| UPenn |  | 1,815 (32%) | 578 (34%) |  |
| **Group** | 7,453 |  |  | 0.3 |
| Alert |  | 2,899 (50%) | 834 (49%) |  |
| Control |  | 2,848 (50%) | 872 (51%) |  |
| **Baseline Risk of Death** | 7,453 | 0.05 (0.03, 0.09) | 0.12 (0.07, 0.19) | <0.001 |
| **Age** | 7,453 | 68 (56, 78) | 66 (56, 76) | <0.001 |
| **Gender** | 7,453 |  |  | 0.050 |
| Female |  | 2,742 (48%) | 768 (45%) |  |
| Male |  | 3,005 (52%) | 938 (55%) |  |
| **Black or African American** | 2,393 | 456 (25%) | 191 (33%) | <0.001 |
| **Teaching Hospital** | 7,453 | 5,747 (100%) | 1,706 (100%) | - |
| **Urban Hospital** | 7,453 | 5,379 (94%) | 1,585 (93%) | 0.3 |
| **ICU** | 7,453 | 523 (9.1%) | 1,357 (80%) | <0.001 |
| **Chronic Heart Failure** | 7,453 | 1,964 (34%) | 407 (24%) | <0.001 |
| **Diabetes** | 7,453 | 1,908 (33%) | 710 (42%) | <0.001 |
| **Malignancy** | 7,453 | 1,320 (23%) | 477 (28%) | <0.001 |
| **Systolic Blood Pressure** | 7,453 | 126 (110, 138) | 120 (106, 133) | <0.001 |
| **Diastolic Blood Pressure** | 7,453 | 71 (64, 78) | 67 (57, 75) | <0.001 |
| **Baseline Creatinine** | 7,453 | 0.94 (0.68, 1.37) | 0.98 (0.67, 1.42) | 0.3 |
| **Creatinine at Randomization** | 7,453 | 1.42 (1.10, 1.91) | 1.50 (1.10, 2.02) | 0.010 |
| **Change in Creatinine** | 7,453 | 0.39 (0.31, 0.52) | 0.40 (0.32, 0.57) | <0.001 |
| **AKI Stage** | 7,453 |  |  | 0.010 |
| 1 |  | 5,294 (92%) | 1,532 (90%) |  |
| 2 |  | 363 (6.3%) | 138 (8.1%) |  |
| 3 |  | 90 (1.6%) | 36 (2.1%) |  |

<sup>1</sup> n (%); Median (IQR)
<sup>2</sup> Pearson's Chi-squared test; Wilcoxon rank sum test

### Provider actions following alerts

Provider actions were associated with alerts. After receiving an alert, providers were more likely to document AKI in the problem list (63% vs. 58%; p<0.001), order IV fluids (39% vs. 37%; p=0.01), and less likely to prescribe NSAIDs (4.3% vs. 5.3%; p=0.043) (Supplementary Tables 5 and 6). The association between provider actions and alerts varied across patient subgroups. Providers were more likely to order IV fluids after an alert for patients with CHF (28% vs. 24%; p=0.004), but not for patients without CHF (47% vs. 45%; p=0.4) (Supplementary Table 7). Providers were more likely to consult nephrology (81% vs. 77%; p=0.002) and start dialysis (77% vs. 73%; p=0.001) after an alert in patients with diastolic blood pressure <70 mmHg, but not in patients with diastolic blood pressure ≥70 mmHg (Supplementary Table 8). For patients on the floor, providers were more likely to order IV fluids (39% vs. 36%; p=0.002) and less likely to prescribe NSAIDs following an alert (4.9% vs. 6.4%; p=0.012) (Supplementary Table 9). For patients in the ICU, there was no difference in the number of orders for IV fluids (39% vs. 39%; p>0.9) or NSAIDs (3.2% vs. 2.9%; p=0.7). Instead, providers were more likely to consult nephrology following an alert (68% vs. 64%; p=0.048). Compared to teaching hospitals, alerts at non-teaching hospitals were more likely to be seen by attending physicians (90% vs. 33%; p<0.001) (Supplementary Table 10).

## DISCUSSION

In this secondary analysis of individual patient data from three randomized controlled trials, we discovered that automated alerts significantly impacted mortality for individual patients. The effect was heterogeneous among patients with different baseline characteristics. Patients with baseline creatinine >1.2 mg/dL had a lower risk of mortality following automated alerts for AKI. Provider actions following alerts may mediate these effects. These results supported our hypothesis that automated alerts can be helpful or harmful, depending on the setting and a patient’s characteristics. In the two external validation trials, optimizing alerts with our model could have theoretically prevented 43 deaths, corresponding to a reduction in the relative risk of all-cause mortality by 6.0% (95% CI: 5.8-6.3%). Assuming that there are 285,000 AKI-related in-hospital deaths per year in the United States,^11^ a 6.0% relative reduction in deaths would correspond to 17,100 deaths per year. The potential implications of these findings are, therefore, considerable.

At the individual level, approximately 77% of patients were predicted to benefit from an alert. The 23% of patients predicted to be harmed from an alert tended to have a higher baseline risk of mortality. As a result, at the population level, the overall effect of alerts on mortality was close to null. This was confirmed by our meta-analysis and is generally consistent with previous studies. One meta-analysis of non-randomized, prospective studies found an association between alerts and decreased mortality (odds ratio [OR]=0.86 [95% CI: 0.75-0.99]; p = 0.040, I^2^= 65.3%; n=5 studies; n=30,791 patients),^12-15^ while another found no mortality benefit [OR=1.05 [95% CI: 0.84-1.31]; n=3 studies; n=3,425 patients; I^2^= 0%].^7^ Reductions in patient mortality could have been attributable to increased provider attention on patients with AKI due to institutional pressures, not from the alert itself.^16,17^ In contrast to the inconsistent results from non-randomized studies, randomized controlled trials have reliably shown no benefit from AKI alerts.^5,8,15^ There was one pilot randomized controlled trial for AKI alerts that was conducted in China but not included in this analysis because of its small sample size.^15^ Mortality at the population-level did not significantly change following the implementation of an alerting system for AKI in an ICU (n=875; 42 deaths [9.0%] in the alert group vs. 33 deaths [8.1%] in the control group; p=0.16). Interactions between alerts and patient characteristics, including baseline creatinine, were not reported in any of these studies due to limitations in either sample size or the inability to test for heterogeneous treatment effects in meta-analyses without individual patient data. Our meta-analysis, which included individual patient data, was able to identify patient and hospital factors that modified the effect of alerts on mortality.

It was difficult to determine the mechanism by which alerts impacted patients’ risk of mortality. AKI is a syndrome that results from a variety of disease processes.^18^ It is possible that alerts may only be beneficial for specific AKI etiologies. For example, if a patient’s AKI is attributable to the administration of a nephrotoxic drug, then alerts may prompt clinicians to stop the offending agent. If an alert is triggered due to a rise in creatinine that is not clinically relevant (false positive),^19^ then prompting a clinician to act on it may lead to iatrogenic harm. Indeed, automated alerts had an observable impact on clinician behavior.^20^ As expected, the impact on care depended on the patient’s phenotype. Patient location within the hospital, a previous diagnosis of CHF, and their blood pressure were all associated with differing effects of alerts. Attending physicians at non-teaching hospitals were more likely to see alerts than attendings at teaching hospitals, which may have been why alerts had a bigger impact on patient care at non-teaching hospitals.

This study has limitations. First, the model may be sensitive to hyperparameter changes. The metric we optimized hyperparameters to, AUC, is a relatively crude measure since iHRs are likely not the most important factor influencing 14-day mortality. We cannot definitively say whether the subgroups affected by alerts in these trials would be affected by alerts in all settings. The external validation provides evidence of generalization across hospitals and alert type (pop-up EHR alerts in the ELAIA-1 trial vs. medication-related EHR alerts in the ELAIA-2 trial vs. text message alerts in the UPenn trial). We cannot say how consistent this effect is in hospitals outside of the Northeast region of the United States. The subgroups identified here were chosen from a pre-specified list of patient characteristics. However, it is possible that considering additional factors would have further refined the patient phenotype expected to be affected by alerts.^21^ It is possible that data leakage may have contaminated the results of our interval validation, since we did not split the ELAIA-1 cohort into “training” and “interval validation” cohorts until after we estimated their baseline risk of death. This could have led to over-optimistic results for the interval validation. However, there was no contamination in the external validations, so readers should put more emphasis on the performance of iHRs in the two external validation cohorts. Finally, the magnitude of alerts on provider actions was relatively small and cannot entirely explain the large effect on mortality. Our analysis of the effect of alerts on provider actions was limited to relatively non-granular data and was at risk of Type 1 error due to multiple testing. Further investigations with more granular data and pre-specified hypotheses are needed.

In conclusion, automated alerts for AKI had a heterogeneous effect on patient mortality. The effect could be predicted from patients’ baseline characteristics. If the two external validation studies used our model to selectively send alerts, there may have been 43 fewer deaths. In our meta-analysis, alerts were associated with a decreased risk of mortality in patients with higher systolic blood pressures but were associated with increased risk of mortality in suburban and non-teaching hospitals. These findings highlight the importance of examining the impact of seemingly benign electronic alerts.

## METHODS

### Trial overviews, study population, and description of the alert intervention

The ELAIA-1 trial was a patient-level, parallel-group, randomized controlled trial conducted in six hospitals in the Yale New Haven Health System in Connecticut and Rhode Island, USA (Supplementary Table 1). The study protocol and results of the primary analysis were previously published.^5,22^ Hospitalized patients aged 18 years or older were enrolled if they met Kidney Disease: Improving Global Outcomes (KDIGO) criteria for AKI: an increase in creatinine by 0.3 mg/dL within 48 hours or a 50% increase over the lowest measured creatinine within the previous week.^23^ Patients were excluded if they were diagnosed with end-stage renal disease or received dialysis within the previous year. For patients randomized to the alert group, an automated, electronic pop-up alert appeared when an intern, resident, fellow, attending physician, nurse practitioner, or physician’s assistant opened their chart. The alert notified the provider that the patient met the criteria for AKI and prompted them to add AKI to the problem list. The alert also contained a link to an AKI order set, which included options for blood and urine testing and kidney imaging.

ELAIA-2 was a follow-up study to ELAIA-1.^9^ There were three key differences between ELAIA-1 and ELAIA-2. First, ELAIA-2 only enrolled patients at the four academic hospitals in the Yale New Haven Health System. It did not enroll patients from the two community hospitals. Second, patients must have been exposed to a non-steroidal anti-inflammatory drug (NSAID), renin angiotensin aldosterone system antagonist, or proton pump inhibitor within 24 hours of enrollment. Third, the intervention prompted clinicians to “consider clinical indication for the following medications,” listed the specific offending agent, and provided a link to the order entry system to discontinue the drug.

The UPenn trial was an investigator-masked, parallel-group, randomized controlled trial.^8^ This trial also used KDIGO criteria to define AKI and used similar inclusion and exclusion criteria (Supplementary Table 1). The alert intervention was a text page that was sent to the covering provider and unit pharmacist’s hospital-provided cell phone. The text contained the patient’s name and room number and said, “Please take appropriate diagnostic and therapeutic measures.”^9^

The institutional review board of Duke University Health System gave ethical approval for this work (#00115914), and all trials data utilized were previously approved. All patient data was obtained from other groups, retrospectivley reviewed, and analyzed. No patients were prospectivley assigned nor treatment altered from our findings.

### Primary outcome

The primary outcome was death within 14 days of randomization. We focused on all-cause mortality rather than the composite outcome of death, progression of AKI, or dialysis because we hypothesized that provider actions prompted by the alerts may improve kidney function at the expense of other organ systems. This theory is supported by results from ELAIA-1, which showed that mortality, not AKI progression or dialysis treatment, was responsible for the heterogeneity of treatment effects across hospitals.^5^ There were no missing outcomes in our data. All analyses were intention-to-treat.

### Data description and pre-processing

We downloaded de-identified ELAIA-1 and ELAIA-2 data from Dryad (https://doi.org/10.5061/dryad.kh189327p), and we obtained data from the UPenn trial from the trial’s principal investigator. Patient characteristics recorded as part of both trials were included and used to predict mortality within 14 days. These included: patient sex, race, age, blood pressure, baseline creatinine, creatinine at the time of randomization, congestive heart failure (CHF) diagnosis, diabetes diagnosis, any malignancy, hospitalization in the intensive care unit, and hospital type (teaching vs. non-teaching and urban vs. non-urban). We used random forest-based imputation with 100 trees to impute missing data based on a matrix of all observed variables in each dataset (*missForest* package).^24^

### Estimation of individual treatment effects

To explore how individual patients experienced the heterogeneous effects of alerts, we created individual prediction models for each patient as described by Oikonomou et al.^25-27^ We calculated individualized hazard ratios (iHRs) based on patient characteristics, where alerts are the independent variable and death within 14 days is the dependent variable, for each patient based on a training dataset. An iHR <1 indicated that alerts were predicted to be protective for that patient, and an iHR >1 indicated that alerts are predicted to be harmful for that patient.

#### Defining iHRs for each patient

First, phenotypic signatures were created from baseline patient characteristics. We created a vector of similarity scores between the index patient and all other patients in the training cohort. Similarity scores were calculated as 1 - Gower’s distance. Gower’s distance quantifies patient dissimilarity on a scale from zero to one. It is calculated as the sum of the difference between two patients’ baseline characteristic values across all variables, divided by the maximum possible difference. This calculation resulted in a similarity matrix with *n* rows (representing *n* patients in the validation cohort) and *p* columns (representing *p* patients in the training cohort). Similarity values were then scaled by raising them to the 10^th^ power and passing them through SoftMax pre-processing and ReLU activation functions (*sigmoid* package).

For each patient, *i* to *n,* in the validation cohort, we fitted a weighted Cox model on patients from the training cohort. We assigned case weights according to the phenotypic similarity between patient *i* in the validation cohort and all patients in the training cohort (*coxphw* package).^28^ The dependent variable was incidence rate of death within 14 days. Time was censored at the time of death, discharge, or 14 days. The independent variables were intervention allocation (alert vs. usual care) and patients’ predicted risk of death at baseline. The Cox model coefficient for alerts was used as the predicted individualized hazard ratio (iHR) of death, or the effect on the estimated risk of death if an alert was sent.

#### Improving precision of iHRs

In order to reduce the unexplained variance in the weighted Cox models, we adjusted iHRs by patients’ baseline risk of death. The predicted risk of death was derived using an ensemble of 100 artificial neural networks (NN) that predicted 14-day mortality based on baseline patient characteristics. Each NN was a multilayer perception that had three hidden layers with 64, 32, and 16 nodes, which was deep enough to model complex interactions between input features.^29^ The NNs were trained to minimize a binary cross entropy loss function with an RMSProp optimizer over 60 epochs, no dropout, and a batch size of 16. We trained the models using a bootstrapping procedure with 100 replicates, each with the same model architecture and hyperparameters but with different initial seeds. Outputs from each NN were averaged to produce each patient’s baseline risk of death over 14 days. For patients discharged from the hospital before 14 days, we assumed they remained alive. The outcome was therefore considered binary (death or no death). Patients in the ELAIA-1 trial were used to train the NN models. These NN models were used to estimate baseline risk of death on patients in the external validation trials (ELAIA-2 and UPenn) without alteration. We used Keras in Python version 3.7.3 for all NN prediction steps.^30^

#### Internal and external validation of estimates of iHRs

Patients in the ELAIA-1 trial were randomly split into training (n=4,221 patients, 70%) and validation cohorts (n=1,809 patients, 30%). We performed five-fold cross-validation on the training cohort for hyperparameter tuning for the model that estimated individualized treatment effects of the alerts. The only hyperparameter we tuned was the relative weight of continuous to categorical variables, used when computing Gower’s distance. The total weight of continuous variables ranged from 10% to 100%, in increments of 10%. Performance was tuned to maximize area under the receiver operating characteristic curve for the model’s ability to predict death within 14 days. Following hyperparameter tuning, we fit the weighted Cox model on all patients in the training cohort, and internally validated it on the patients in the holdout validation cohort. For the external validations, iHRs for patients in the ELAIA-2 and UPenn trials were computed using models fit on all patients from the ELAIA-1 trial. For each patient *i* to *n* in the ELAIA-2 and UPenn trials, iHRs were calculated using the following steps:

**Step 1.** Estimate patient *i*’s baseline risk of death using NNs trained on patients in the ELAIA-1 trial.

**Step 2.** Calculate patient *i*’s similarity (Gower’s distance) to each patient in the ELAIA-1 trial.

**Step 3.** Fit a weighted Cox model on patients in the ELAIA-1 trial, weighting each observation (patient) according to their similarity to patient *i* (calculated in Step 2). R code:

*cox_model_fit <-coxphw(Surv(time, death14days) ~ alert + baseline_risk_of_death, data = ELAIA1_trial_data, caseweights = similarity_scores)*

Repeating Steps 1-3 for all patients *i* to *n* in the two external validation trials allowed us to compare the observed outcomes against their predicted benefit from an alert. We compared survival between patients receiving beneficial and harmful alerts. The observed treatment effect of alerts on mortality was calculated using Cox proportional hazards models. Interactions between the predicted log-odds of alert benefit, the intervention allocation (alert vs. usual care), and 14-day mortality were calculated with an adjustment for baseline risk of mortality (predicted using NNs as described above). The baseline risk of mortality was included as a covariate in the Cox model. To visualize survival in patients who received beneficial or harmful alerts, we dichotomized patients with iHR<1 and iHR>1, respectively, and plotted Kaplan-Meier curves. There were no patients with iHR = 1.

#### Patient characteristics’ contribution to iHR

Patient baseline characteristics were ultimately used to estimate the effect of alerts on their risk of mortality. To understand how patient characteristics contributed to the prediction of individualized benefit from an alert, we trained an XGBoost model to indirectly predict a patient’s iHR based on their baseline clinical characteristics.^31^ XGBoost is a gradient-boosted tree-based machine learning algorithm. It was chosen because it performs well on tabular data and can model complex, non-linear interactions between predictors. We trained the XGBoost model on ELAIA-1 and applied it to patients from the UPenn trial. Since this was done for explanatory purposes only, we were not concerned about recycling data that we previously used to calculate iHRs with the weighted Cox models. We used Bayesian optimization for XGBoost hyperparameter tuning (*ParBayesianOptimization* package). Hyperparameters included: learning rate (eta range: 0.1 to 0.3), minimum loss reduction need to split a node (gamma range: 0 to 100), max depth of the tree (3 to 10), minimum child weight (range: 1 to 10), and subsample ratio used for each training instance (range: 0.5 to 1). We set the maximum number of boosting iterations to 500, with early stopping if there was no improvement in performance for 20 rounds. Optimal hyperparameters were selected using five-fold cross-validation. We initialized the Gaussian process with 20 points and ran the Bayesian search for five iterations. After training, we used SHAP (SHapley Additive exPlanations) values to quantify a specific feature value’s contribution to the predicted treatment effect.^32^ SHAP values were summarized using a beeswarm plot (*xgboost* package).

### Individual patient data meta-analysis

We conducted an individual patient data meta-analysis to investigate the linear relationship between patient characteristics and the effect of alerts on mortality. This complimented our prediction of iHRs, which captured non-linear relationships between patient characteristics and the effect of alerts on mortality. For the meta-analysis, we followed the Preferred Reporting Items for a Systematic Review and Meta-analysis of Individual Participant Data (PRISMA-IPD) guidelines for reporting.^33^ The attached MEDLINE search was conducted for studies from 2015 onward in January, 2024. Randomized control trials investigating automated electronic alerts for AKI with a primary outcome of death written in English were included. Reviews, meta-analyses, other secondary analysis were excluded. Risk of bias was assessed using Version 2 of the Cochrane Risk-of-Bias Assessment tool.^10^ We conducted a search to ensure that no other randomized controlled trials were conducted that met our inclusion criteria. The analyses reported here were considered exploratory and were not pre-specified in a registered statistical analysis plan.

De-identified patient data were assumed to be non-duplicative within and across trials. We verified that patient data were consistent with previous publications. Covariates and possible interaction terms were selected from the previously published list of covariates in the Electronic Alerts for Acute Kidney Injury Amelioration (ELAIA-1) trial.^5^ Patient characteristics recorded as part of both trials were included and used to predict mortality within 14 days. These included: patient sex, race, age, blood pressure, baseline creatinine, creatinine at the time of randomization, congestive heart failure (CHF) diagnosis, diabetes diagnosis, any malignancy, hospitalization in the intensive care unit, and hospital type (teaching vs. non-teaching and urban vs. non-urban). Continuous variables (age, creatinine, and blood pressure) were treated as such when testing for treatment interactions. For visualization purposes, variables were categorized into groups and shown in a forest plot. Cut points were chosen based on the ELAIA-1 trial publication.^5^ The original trials did not provide lists of specific ICD codes that they used to define each medical comorbidity, so we merged comorbidity labels across trials by name. The trials also did not specify a time element associated with them (e.g. malignancy within the last five years).

We used a multivariable Cox proportional hazards model, stratified by trial, to estimate the hazard of mortality using the patient characteristics mentioned above and treatment interactions as independent variables. Using a Cox model, rather than the modified Poisson generalized estimating equations used in the ELAIA-1 trial, allowed us to account for censoring. A single model was fit with all interaction terms and included data from all three trials. Because ELAIA-2 did not include data about patient race, we were unable to model the interaction of that variable with alerts. We included interaction terms for each hospital, under the assumption that alerts may have different effects at different hospitals. All effect estimates were reported using adjusted hazard ratios (HR) with the corresponding 95% confidence intervals (CI) comparing patients randomized to the alert group against those in the control group. Time (days) was censored at death, discharge, or 14 days, whichever came first. We used Schoenfeld residuals to test whether the proportional hazards assumption was satisfied (“cox.zph” function from the *survival* package).^34^ P<0.05 for ICU status and systolic blood pressure variables, indicating violation of the proportional hazards assumption. Stratifying the model by ICU status eliminated the violation, but the model fit, as measured by concordance, significantly decreased. Therefore, we chose to leave the model as is. We assessed heterogeneity using Higgins & Thompson’s I^2^ statistic, defined as the percentage of variability in the effect sizes that is not caused by sampling (“rma” function from the *metafor* package). It quantifies between-study heterogeneity and is directly based on Cochran’s Q but is more stable for small numbers of trials.^35,36^ Treatment interactions were assumed to be statistically significant if two-sided p-values were less than 0.05. We did not adjust p-values for multiple comparisons. All statistical analyses were performed using R version 4.4.1.

### Provider actions following alerts

We assumed that any effect that alerts had on patient outcomes must have been mediated by provider actions following those alerts. To investigate this, we compared the frequency of AKI documentation, orders for IV fluids, urinalyses, ACE/ARBs, NSAIDs, aminoglycosides, dialysis, and nephrology consults between patients randomized to the alert or usual care groups. Data were taken from the ELAIA-1 and UPenn trials. The ELAIA-2 trial was not included. Differences in frequencies were compared using Pearson’s Chi-squared test or Fisher’s exact test, and difference in continuous measures were compared using Wilcoxon rank sum test. We stratified patients by the following groups: trial, diagnosis of CHF, diastolic blood pressure <70 mmHg, hospital teaching status, and ICU status.

### Exploratory analysis of patient phenotype heterogeneity

To investigate the heterogeneity of patient phenotypes, we quantified the relative impact of patient characteristics on their risk of death. We trained an ensemble of NNs to predict the risk of death at 14 days using all 53 baseline variables there were available prior to randomization in the ELAIA-1 trial, and SHAP values to estimate the impact that each variable had on a patient’s predicted risk of death. Patient information were used as the input to a NN with three hidden layers with 64, 32, and 16 nodes, deep enough to model complex interactions between input features that conventional regression techniques do not capture.^29^ The NN was trained to minimize a binary cross entropy loss function with an rms-prop optimizer over three epochs and a batch size of 16. Patients were randomly split 70:30 into training and validation datasets. Patients in the training dataset were used to train the NN while patients in the validation dataset were used to 1) estimate model performance and 2) interrogate which features drove the model’s predictions. We used Keras to build the NNs and the SHAP package to explain predictions in Python version 3.7.3.^30,32^

An ensemble of NNs was built using a bootstrapping procedure with 50 replicates, each with the same model architecture and hyperparameters, but different initial seeds. Training and validation cohorts were randomly chosen for each bootstrapped iteration, resulting in 50 distinct NNs. Patients were never included in both the training and validation datasets in the same iteration. Model performance was reported as the area under the receiver operator characteristic curve (AUC) on the validation datasets from the ensemble predictions from all 50 NNs.^37^ Confidence intervals were computed using DeLong’s method.^38^ Since each patient had SHAP values from multiple NNs, values for each feature were averaged.^39,40^

The NN estimated each patient’s risk of death, and SHAP values were used to explain the predictions, giving the ability to quantify the relative impact of a specific feature on a patient’s predicted risk of death.^32,41^ When reporting SHAP values, one-hot features for hospitals 1 through 6 were merged into one category called “location”, and “ICU” and “ward” were also combined into one category.^41^

Patients were visualized using T-Distributed Stochastic Neighbor Embedding (t-SNE), a dimensionality reduction technique.^42^ The input to the t-SNE algorithm was the SHAP values (relative contributions) of each feature on patients’ predicted risk of death. This allowed patients whose risk of death was attributable to similar characteristics to be visualized near each other in a two-dimensional space. Default hyperparameters for the *Rtsne* package were used.^43^ t-SNE plots were color coded according to patients’ baseline characteristics (Figure 1 and Supplemental Figure 3) or the SHAP values for the features available at baseline (Supplemental Figure 4).

## Supporting information

Supplementary Results

## Data Availability

We downloaded de-identified ELAIA-1 and ELAIA-2 data from Dryad (https://doi.org/10.5061/dryad.kh189327p), and we obtained data from the UPenn trial from the trial's principal investigator.

https://github.com/wisselbd/AKI_Alerts

## Data availability

All of the data used in these analyses are publicly available on DRYAD.

## Code availability

The analytical code used to develop the deep neural networks in Python and the subsequent analyses performed in R will be made available on GitHub upon publication. The trained XGBoost model is available on GitHub (https://github.com/wisselbd/AKI_Alerts). All statistical analyses were performed using R (v4.4.1). The neural networks utilized Python (v3.7.3) and Keras (v2.9.0). The following packaged were utilized in R: missForest (v1.4.0), sigmoid (v1.4.0), coxphw (v4.0.1), ParBayesianOptimization (v1.2.6), xgboost (v1.6.0.1), survival (v3.2.1), metafor (v4.6.0), rtsne (v0.17.0).

## Acknowledgments

We would like to sincerely thank the ELAIA and UPenn study teams for graciously releasing the trial data. This work was partially supported by a grant from the National Institutes of Health (F31 NS115447) to Benjamin Wissel.

## Author contributions

All contributors who met ICJME criteria for authorship were included. B.D.W. and Z.P. conceived of the idea for the analysis, performed the statistical analyses, and wrote the first draft of the manuscript. E.G. performed statistical analyses, helped with revisions and approved the final submission. B.B., T.J.Z., I.S.K., S.L.G, and J.W.D. provided significant intellectual feedback and helped with revisions. All authors have read and approved the final submission. The corresponding author attests to the integrity of the analysis, accepts full responsibility for the overall work, has access to the data, and controlled the decision to publish.

