## Supplementary Results for "Heterogeneous Effect of Automated Alerts on Mortality"

**SUPPLEMENTARY TABLES AND FIGURES**

**Supplementary Table 1.** Characteristics of alert and control groups for all participants in both included studies.

| Characteristic | Overall, N = 13,483*^1^* | Alert, N = 6,792*^1^* | Control, N = 6,691*^1^* | p-value*^2^* |
| --- | --- | --- | --- | --- |
| Study |  |  |  | 0.8 |
| ELAIA-1 | 6,030 (45%) | 3,059 (45%) | 2,971 (44%) |  |
| ELAIA-2 | 5,060 (38%) | 2,532 (37%) | 2,528 (38%) |  |
| UPenn | 2,393 (18%) | 1,201 (18%) | 1,192 (18%) |  |
| Death | 1,250 (9.3%) | 618 (9.1%) | 632 (9.4%) | 0.5 |
| Age | 69 (58, 80) | 69 (57, 80) | 69 (58, 79) | >0.9 |
| Sex |  |  |  | 0.5 |
| Female | 6,392 (47%) | 3,199 (47%) | 3,193 (48%) |  |
| Male | 7,091 (53%) | 3,593 (53%) | 3,498 (52%) |  |
| Black or African American | 1,593 (19%) | 808 (19%) | 785 (19%) | 0.9 |
| Teaching Hospital | 12,718 (94%) | 6,413 (94%) | 6,305 (94%) | 0.6 |
| Urban Hospital | 11,662 (86%) | 5,890 (87%) | 5,772 (86%) | 0.4 |
| ICU | 3,803 (28%) | 1,887 (28%) | 1,916 (29%) | 0.3 |
| Chronic Heart Failure | 5,029 (37%) | 2,565 (38%) | 2,464 (37%) | 0.3 |
| Diabetes | 5,102 (38%) | 2,607 (38%) | 2,495 (37%) | 0.2 |
| Malignancy | 2,728 (20%) | 1,360 (20%) | 1,368 (20%) | 0.5 |
| Systolic Blood Pressure | 121 (107, 136) | 122 (107, 137) | 121 (106, 134) | <0.001 |
| Diastolic Blood Pressure | 69 (60, 77) | 69 (60, 77) | 69 (60, 77) | 0.4 |
| Baseline Creatinine | 0.98 (0.70, 1.40) | 0.99 (0.70, 1.40) | 0.98 (0.69, 1.40) | 0.4 |
| Creatinine at Randomization | 1.48 (1.12, 1.98) | 1.49 (1.13, 1.99) | 1.47 (1.10, 1.98) | 0.5 |
| Change in Creatinine | 0.40 (0.32, 0.54) | 0.40 (0.32, 0.54) | 0.40 (0.32, 0.55) | 0.7 |

*^1^* n (%); Median (IQR)

*^2^* Pearson's Chi-squared test; Wilcoxon rank sum test

**Supplementary Table 2.** Characteristics of alert and control groups for participants treated in non-teaching hospitals.

| Characteristic | Overall, N = 765*^1^* | Alert, N = 379*^1^* | Control, N = 386*^1^* | p-value*^2^* |
| --- | --- | --- | --- | --- |
| Study |  |  |  |  |
| ELAIA-1 | 765 (100%) | 379 (100%) | 386 (100%) |  |
| Death | 92 (12%) | 59 (16%) | 33 (8.5%) | 0.003 |
| Age | 74 (63, 85) | 74 (63, 85) | 73 (64, 84) | 0.8 |
| Sex |  |  |  | 0.3 |
| Female | 370 (48%) | 190 (50%) | 180 (47%) |  |
| Male | 395 (52%) | 189 (50%) | 206 (53%) |  |
| Black or African American | 44 (5.8%) | 24 (6.3%) | 20 (5.2%) | 0.5 |
| Urban Hospital | 0 (0%) | 0 (0%) | 0 (0%) |  |
| ICU | 391 (51%) | 206 (54%) | 185 (48%) | 0.075 |
| Chronic Heart Failure | 387 (51%) | 200 (53%) | 187 (48%) | 0.2 |
| Diabetes | 323 (42%) | 157 (41%) | 166 (43%) | 0.7 |
| Malignancy | 90 (12%) | 45 (12%) | 45 (12%) | >0.9 |
| Systolic Blood Pressure | 118 (105, 134) | 118 (104, 134) | 118 (105, 135) | 0.8 |
| Diastolic Blood Pressure | 65 (56, 76) | 65 (56, 76) | 65 (56, 76) | 0.7 |
| Baseline Creatinine | 1.16 (0.80, 1.63) | 1.14 (0.80, 1.55) | 1.21 (0.82, 1.70) | 0.10 |
| Creatinine at Randomization | 1.70 (1.26, 2.23) | 1.64 (1.25, 2.17) | 1.75 (1.27, 2.32) | 0.11 |
| Change in Creatinine | 0.44 (0.34, 0.64) | 0.44 (0.34, 0.63) | 0.44 (0.34, 0.64) | 0.9 |

*^1^* n (%); Median (IQR)

*^2^* Pearson's Chi-squared test; Wilcoxon rank sum test

**Supplementary Table 3.** Characteristics of alert and control groups for participants with diastolic blood pressure < 70mmHg.

| **Characteristic** | **Overall**, N = 7,108*^1^* | **Alert**, N = 3,574*^1^* | **Control**, N = 3,534*^1^* | **p-value***^2^* |
| --- | --- | --- | --- | --- |
| **Age** | 73 (62, 82) | 73 (62, 82) | 73 (62, 82) | 0.6 |
| **Gender** |  |  |  | >0.9 |
| Female | 3,444 (48%) | 1,733 (48%) | 1,711 (48%) |  |
| Male | 3,664 (52%) | 1,841 (52%) | 1,823 (52%) |  |
| **Black or African American** | 523 (12%) | 261 (12%) | 262 (13%) | 0.6 |
| **Teaching Hospital** | 6,642 (93%) | 3,340 (93%) | 3,302 (93%) | >0.9 |
| **Urban Hospital** | 5,957 (84%) | 2,999 (84%) | 2,958 (84%) | 0.8 |
| **ICU** | 2,458 (35%) | 1,233 (34%) | 1,225 (35%) | 0.9 |
| **Chronic Heart Failure** | 2,953 (42%) | 1,520 (43%) | 1,433 (41%) | 0.090 |
| **Diabetes** | 2,838 (40%) | 1,452 (41%) | 1,386 (39%) | 0.2 |
| **Malignancy** | 1,400 (20%) | 687 (19%) | 713 (20%) | 0.3 |
| **Systolic Blood Pressure** | 110 (99, 123) | 110 (99, 123) | 110 (99, 123) | 0.4 |
| **Diastolic Blood Pressure** | 61 (54, 65) | 61 (54, 65) | 61 (54, 66) | >0.9 |
| **Baseline Creatinine** | 1.00 (0.72, 1.43) | 1.00 (0.71, 1.44) | 1.00 (0.72, 1.43) | >0.9 |
| **Creatinine at Randomization** | 1.53 (1.18, 2.02) | 1.53 (1.18, 2.00) | 1.54 (1.19, 2.04) | 0.4 |
| **Change in Creatinine** | 0.41 (0.33, 0.58) | 0.41 (0.33, 0.57) | 0.41 (0.33, 0.60) | 0.2 |

*^1^* n (%); Median (IQR)

*^2^* Pearson's Chi-squared test; Wilcoxon rank sum test

**Supplementary Table 4.** Characteristics of alert and control groups for participants with baseline creatinine greater than 1.2 mg/dL.

| **Characteristic** | **Overall**, N = 4,581*^1^* | **Alert**, N = 2,303*^1^* | **Control**, N = 2,278*^1^* | **p-value***^2^* |
| --- | --- | --- | --- | --- |
| **Age** | 73 (62, 82) | 73 (61, 82) | 73 (62, 83) | 0.2 |
| **Gender** |  |  |  | 0.4 |
| Female | 1,723 (38%) | 879 (38%) | 844 (37%) |  |
| Male | 2,858 (62%) | 1,424 (62%) | 1,434 (63%) |  |
| **Black or African American** | 632 (22%) | 319 (22%) | 313 (21%) | 0.7 |
| **Teaching Hospital** | 4,216 (92%) | 2,132 (93%) | 2,084 (91%) | 0.2 |
| **Urban Hospital** | 3,874 (85%) | 1,968 (85%) | 1,906 (84%) | 0.095 |
| **ICU** | 1,295 (28%) | 639 (28%) | 656 (29%) | 0.4 |
| **Chronic Heart Failure** | 2,402 (52%) | 1,200 (52%) | 1,202 (53%) | 0.7 |
| **Diabetes** | 2,238 (49%) | 1,135 (49%) | 1,103 (48%) | 0.6 |
| **Malignancy** | 833 (18%) | 415 (18%) | 418 (18%) | 0.8 |
| **Systolic Blood Pressure** | 123 (108, 139) | 124 (108, 141) | 122 (108, 137) | 0.069 |
| **Diastolic Blood Pressure** | 68 (59, 76) | 68 (59, 76) | 68 (60, 76) | >0.9 |
| **Baseline Creatinine** | 1.65 (1.40, 2.13) | 1.65 (1.40, 2.12) | 1.65 (1.40, 2.14) | 0.8 |
| **Creatinine at Randomization** | 2.18 (1.86, 2.71) | 2.16 (1.86, 2.70) | 2.19 (1.86, 2.73) | 0.5 |
| **Change in Creatinine** | 0.41 (0.34, 0.58) | 0.41 (0.33, 0.57) | 0.42 (0.34, 0.59) | 0.048 |

*^1^* n (%); Median (IQR)

*^2^* Pearson's Chi-squared test; Wilcoxon rank sum test

**Supplementary Table 5.** Provider actions following randomization. Intravenous (IV) fluid, urinalysis, and nephrotoxic drug orders were recorded within 24 hours of randomization. Acute kidney injury (AKI) documentation, nephrology consults, and orders for dialysis were recorded any time prior to discharge. ACE: angiotensin converting enzyme; ARB: angiotensin receptor blocker; NSAID: non-steroidal anti-inflammatory drug.

|  | Alert, N = 4,260*^1^* | Usual Care, N = 4,163*^1^* | p-value*^2^* |
| --- | --- | --- | --- |
| AKI Documentation | 2,686 (63%) | 2,402 (58%) | <0.001 |
| IV Fluids | 1,674 (39%) | 1,522 (37%) | 0.010 |
| IV Fluid Bolus | 442 (10%) | 375 (9.0%) | 0.034 |
| Urinalysis | 711 (17%) | 638 (15%) | 0.088 |
| Nephrotoxic Drug(s) | 801 (19%) | 829 (20%) | 0.2 |
| ACE/ARBs | 593 (14%) | 578 (14%) | >0.9 |
| NSAIDs | 185 (4.3%) | 220 (5.3%) | 0.043 |
| Aminoglycosides | 49 (1.2%) | 67 (1.6%) | 0.071 |
| Nephrology Consult | 2,943 (69%) | 2,834 (68%) | 0.3 |
| Dialysis | 2,771 (65%) | 2,659 (64%) | 0.3 |

*^1^* n (%)

*^2^* Pearson's Chi-squared test

**Supplementary Table 6.** Provider actions following randomization in the ELAIA-1 and UPenn trials. Intravenous (IV) fluid, urinalysis, and nephrotoxic drug orders were recorded within 24 hours of randomization. Acute kidney injury (AKI) documentation, nephrology consults, and orders for dialysis were recorded any time prior to discharge. ACE: angiotensin converting enzyme; ARB: angiotensin receptor blocker; NSAID: non-steroidal anti-inflammatory drug.

|  | ELAIA-1 | | | UPenn | | |
| --- | --- | --- | --- | --- | --- | --- |
|  | **Alert, N = 3,059*^1^*** | **Usual Care, N = 2,971*^1^*** | **p-value*^2^*** | **Alert, N = 1,201*^1^*** | **Usual Care, N = 1,192*^1^*** | **p-value*^2^*** |
| AKI Documentation | 2,141 (70%) | 1,871 (63%) | <0.001 | 545 (45%) | 531 (45%) | 0.7 |
| IV Fluids | 1,174 (38%) | 1,034 (35%) | 0.004 | 500 (42%) | 488 (41%) | 0.7 |
| IV Fluid Bolus | 397 (13%) | 339 (11%) | 0.063 | 45 (3.7%) | 36 (3.0%) | 0.3 |
| Urinalysis | 519 (17%) | 444 (15%) | 0.032 | 192 (16%) | 194 (16%) | 0.8 |
| Nephrotoxic Drug(s) | 563 (18%) | 578 (19%) | 0.3 | 238 (20%) | 251 (21%) | 0.5 |
| ACE/ARBs | 424 (14%) | 425 (14%) | 0.6 | 169 (14%) | 153 (13%) | 0.4 |
| NSAIDs | 144 (4.7%) | 166 (5.6%) | 0.12 | 41 (3.4%) | 54 (4.5%) | 0.2 |
| Aminoglycosides | 14 (0.5%) | 19 (0.6%) | 0.3 | 35 (2.9%) | 48 (4.0%) | 0.14 |
| Nephrology Consult | 2,814 (92%) | 2,722 (92%) | 0.6 | 129 (11%) | 112 (9.4%) | 0.3 |
| Dialysis | 2,673 (87%) | 2,578 (87%) | 0.5 | 98 (8.2%) | 81 (6.8%) | 0.2 |

*^1^* n (%)

*^2^* Pearson's Chi-squared test

**Supplementary Table 7.** Provider actions following randomization stratified by patient diagnosis of congestive heart failure (CHF). Intravenous (IV) fluid, urinalysis, and nephrotoxic drug orders were recorded within 24 hours of randomization. Acute kidney injury (AKI) documentation, nephrology consults, and orders for dialysis were recorded any time prior to discharge. ACE: angiotensin converting enzyme; ARB: angiotensin receptor blocker; NSAID: non-steroidal anti-inflammatory drug.

|  | CHF | | | No CHF | | |
| --- | --- | --- | --- | --- | --- | --- |
|  | **Alert**, N = 1,738*^1^* | **Usual Care**, N = 1,680*^1^* | **p-value***^2^* | **Alert**, N = 2,522*^1^* | **Usual Care**, N = 2,483*^1^* | **p-value***^2^* |
| AKI Documentation | 1,266 (73%) | 1,152 (69%) | 0.006 | 1,420 (56%) | 1,250 (50%) | <0.001 |
| IV Fluids | 495 (28%) | 406 (24%) | 0.004 | 1,179 (47%) | 1,116 (45%) | 0.2 |
| IV Fluid Bolus | 148 (8.5%) | 117 (7.0%) | 0.090 | 294 (12%) | 258 (10%) | 0.2 |
| Urinalysis | 289 (17%) | 244 (15%) | 0.090 | 422 (17%) | 394 (16%) | 0.4 |
| Nephrotoxic Drug(s) | 318 (18%) | 329 (20%) | 0.3 | 483 (19%) | 500 (20%) | 0.4 |
| ACE/ARBs | 282 (16%) | 302 (18%) | 0.2 | 311 (12%) | 276 (11%) | 0.2 |
| NSAIDs | 30 (1.7%) | 23 (1.4%) | 0.4 | 155 (6.1%) | 197 (7.9%) | 0.013 |
| Aminoglycosides | 13 (0.7%) | 12 (0.7%) | >0.9 | 36 (1.4%) | 55 (2.2%) | 0.037 |
| Nephrology Consult | 1,285 (74%) | 1,233 (73%) | 0.7 | 1,658 (66%) | 1,601 (64%) | 0.3 |
| Dialysis | 1,206 (69%) | 1,141 (68%) | 0.4 | 1,565 (62%) | 1,518 (61%) | 0.5 |

*^1^* n (%)

*^2^* Pearson's Chi-squared test

**Supplementary Table 8.** Provider actions following randomization stratified by diastolic blood pressure. Intravenous (IV) fluid, urinalysis, and nephrotoxic drug orders were recorded within 24 hours of randomization. Acute kidney injury (AKI) documentation, nephrology consults, and orders for dialysis were recorded any time prior to discharge. ACE: angiotensin converting enzyme; ARB: angiotensin receptor blocker; NSAID: non-steroidal anti-inflammatory drug.

|  | Diastolic BP <70 mmHg | | | Diastolic BP ≥70 mmHg | | |
| --- | --- | --- | --- | --- | --- | --- |
|  | **Alert**, N = 2,133*^1^* | **Usual Care**, N = 2,094*^1^* | **p-value***^2^* | **Alert**, N = 2,127*^1^* | **Usual Care**, N = 2,069*^1^* | **p-value***^2^* |
| AKI Documentation | 1,480 (69%) | 1,343 (64%) | <0.001 | 1,206 (57%) | 1,059 (51%) | <0.001 |
| IV Fluids | 871 (41%) | 790 (38%) | 0.039 | 803 (38%) | 732 (35%) | 0.11 |
| IV Fluid Bolus | 272 (13%) | 228 (11%) | 0.061 | 170 (8.0%) | 147 (7.1%) | 0.3 |
| Urinalysis | 376 (18%) | 328 (16%) | 0.087 | 335 (16%) | 310 (15%) | 0.5 |
| Nephrotoxic Drug(s) | 348 (16%) | 371 (18%) | 0.2 | 453 (21%) | 458 (22%) | 0.5 |
| ACE/ARBs | 252 (12%) | 266 (13%) | 0.4 | 341 (16%) | 312 (15%) | 0.4 |
| NSAIDs | 87 (4.1%) | 100 (4.8%) | 0.3 | 98 (4.6%) | 120 (5.8%) | 0.082 |
| Aminoglycosides | 19 (0.9%) | 22 (1.1%) | 0.6 | 30 (1.4%) | 45 (2.2%) | 0.062 |
| Nephrology Consult | 1,730 (81%) | 1,619 (77%) | 0.002 | 1,213 (57%) | 1,215 (59%) | 0.3 |
| Dialysis | 1,644 (77%) | 1,524 (73%) | 0.001 | 1,127 (53%) | 1,135 (55%) | 0.2 |

*^1^* n (%)

*^2^* Pearson's Chi-squared test

**Supplementary Table 9.** Provider actions following randomization stratified by patient location within the hospital (intensive care unit [ICU] vs. floor). Intravenous (IV) fluid, urinalysis, and nephrotoxic drug orders were recorded within 24 hours of randomization. Acute kidney injury (AKI) documentation, nephrology consults, and orders for dialysis were recorded any time prior to discharge. ACE: angiotensin converting enzyme; ARB: angiotensin receptor blocker; NSAID: non-steroidal anti-inflammatory drug.

|  | Floor | | | ICU | | |
| --- | --- | --- | --- | --- | --- | --- |
|  | **Alert**, N = 2,933*^1^* | **Usual Care**, N = 2,845*^1^* | **p-value***^2^* | **Alert**, N = 1,327*^1^* | **Usual Care**, N = 1,318*^1^* | **p-value***^2^* |
| AKI Documentation | 1,786 (61%) | 1,543 (54%) | <0.001 | 900 (68%) | 859 (65%) | 0.15 |
| IV Fluids | 1,153 (39%) | 1,005 (35%) | 0.002 | 521 (39%) | 517 (39%) | >0.9 |
| IV Fluid Bolus | 286 (9.8%) | 219 (7.7%) | 0.006 | 156 (12%) | 156 (12%) | >0.9 |
| Urinalysis | 453 (15%) | 393 (14%) | 0.080 | 258 (19%) | 245 (19%) | 0.6 |
| Nephrotoxic Drug(s) | 648 (22%) | 663 (23%) | 0.3 | 153 (12%) | 166 (13%) | 0.4 |
| ACE/ARBs | 502 (17%) | 480 (17%) | 0.8 | 91 (6.9%) | 98 (7.4%) | 0.6 |
| NSAIDs | 143 (4.9%) | 182 (6.4%) | 0.012 | 42 (3.2%) | 38 (2.9%) | 0.7 |
| Aminoglycosides | 26 (0.9%) | 32 (1.1%) | 0.4 | 23 (1.7%) | 35 (2.7%) | 0.11 |
| Nephrology Consult | 2,040 (70%) | 1,985 (70%) | 0.9 | 903 (68%) | 849 (64%) | 0.048 |
| Dialysis | 1,933 (66%) | 1,868 (66%) | 0.8 | 838 (63%) | 791 (60%) | 0.10 |

*^1^* n (%)

*^2^* Pearson's Chi-squared test

**Supplementary Table 10.** Provider actions following randomization stratified by hospital type (teaching vs. non-teaching). Intravenous (IV) fluid, urinalysis, and nephrotoxic drug orders were recorded within 24 hours of randomization. Acute kidney injury (AKI) documentation, nephrology consults, and orders for dialysis were recorded any time prior to discharge. ACE: angiotensin converting enzyme; ARB: angiotensin receptor blocker; NSAID: non-steroidal anti-inflammatory drug.

|  | Non-Teaching Hospital | | | Teaching Hospital | | |
| --- | --- | --- | --- | --- | --- | --- |
|  | **Alert**, N = 379*^1^* | **Usual Care**, N = 386*^1^* | **p-value***^2^* | **Alert**, N = 3,881*^1^* | **Usual Care**, N = 3,777*^1^* | **p-value***^3^* |
| AKI Documentation | 254 (67%) | 243 (63%) | 0.2 | 2,432 (63%) | 2,159 (57%) | <0.001 |
| IV Fluids | 149 (39%) | 138 (36%) | 0.3 | 1,525 (39%) | 1,384 (37%) | 0.017 |
| IV Fluid Bolus | 22 (5.8%) | 25 (6.5%) | 0.7 | 420 (11%) | 350 (9.3%) | 0.024 |
| Urinalysis | 46 (12%) | 45 (12%) | 0.8 | 665 (17%) | 593 (16%) | 0.090 |
| Nephrotoxic Drug(s) | 83 (22%) | 92 (24%) | 0.5 | 718 (19%) | 737 (20%) | 0.3 |
| ACE/ARBs | 65 (17%) | 72 (19%) | 0.6 | 528 (14%) | 506 (13%) | 0.8 |
| NSAIDs | 23 (6.1%) | 23 (6.0%) | >0.9 | 162 (4.2%) | 197 (5.2%) | 0.031 |
| Aminoglycosides | 0 (0%) | 1 (0.3%) | >0.9 | 49 (1.3%) | 66 (1.7%) | 0.081 |
| Nephrology Consult | 366 (97%) | 365 (95%) | 0.2 | 2,577 (66%) | 2,469 (65%) | 0.3 |
| Dialysis | 349 (92%) | 347 (90%) | 0.3 | 2,422 (62%) | 2,312 (61%) | 0.3 |

*^1^* n (%)

*^2^* Pearson's Chi-squared test; Fisher’s exact test

*^3^* Pearson's Chi-squared test

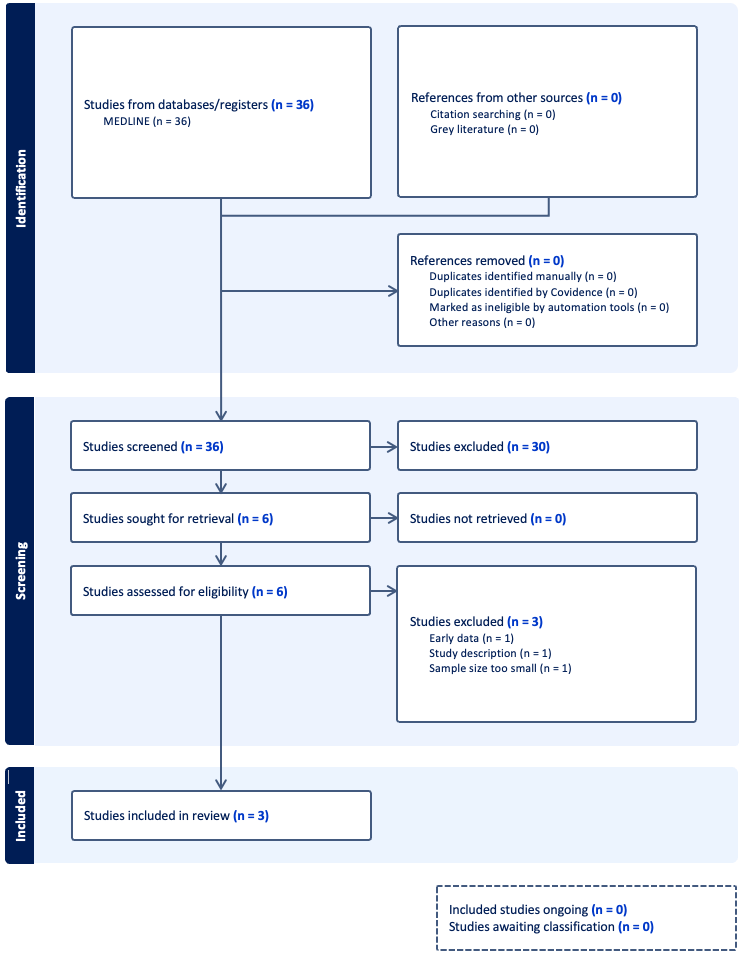

**Supplementary Figure 1. PRISMA Flow Diagram. Studies underwent abstract screening and full text screening prior to inclusion in the study.**

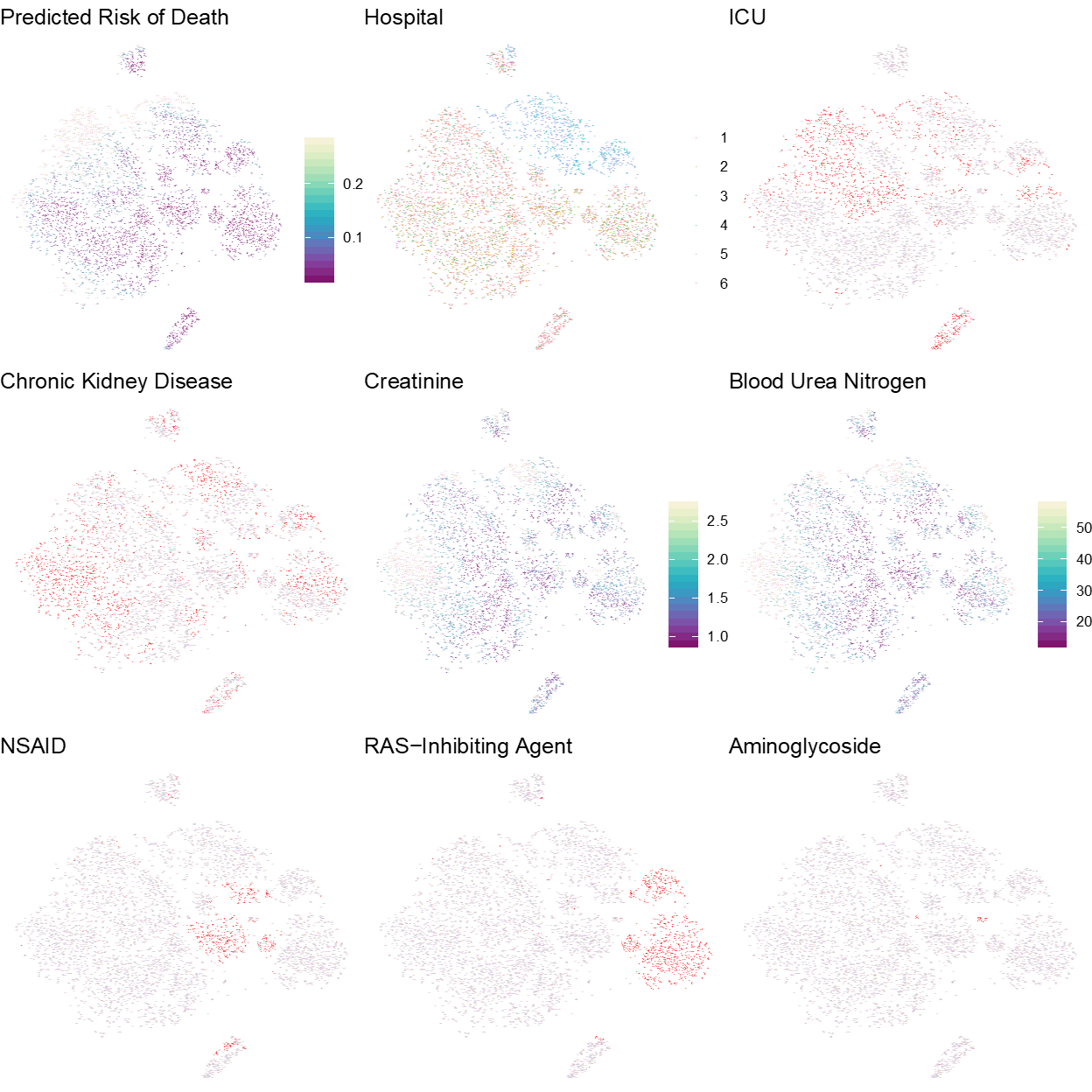

**Supplementary Figure 2**. Unsupervised t-SNE visualization of features that contributed to patients’ predicted risk of death (ELAIA-1 trial; n=6,030 patients). Each dot represents one patient. Each panel includes the same set of patients with different color codes. Binary features were colored red when present and grey when absent. “Predicted Risk of Death” (top left) was the deep neural network’s predicted risk of mortality within 14 days of randomization. Patients in Hospitals 4 and 5 clustered together (top, middle). Patients with chronic kidney disease tended to have higher creatinine and blood urea nitrogen levels. Patients exposed to nephrotoxic drugs in the 24 hours prior to randomization (bottom row) clustered together. ICU: intensive care unit; NSAID: nonsteroidal anti-inflammatory drug; RAS: renin-angiotensin system.

**Supplementary Figure 3.** t-SNE plot colored by patient characteristics. Each dot represents one patient from the ELAIA-1 trial.

**Supplementary Figure 4.** t-SNE plot colored by SHAP values for patient characteristics. Each dot represents one patient from the ELAIA-1 trial. Dots were colored grey when the corresponding SHAP value was at the population median, red when in the 10^th^ percentile (i.e. feature value decreased the model’s predicted risk of death), and blue when in the 90^th^ percentile (i.e. feature value increased the model’s predicted risk of death).

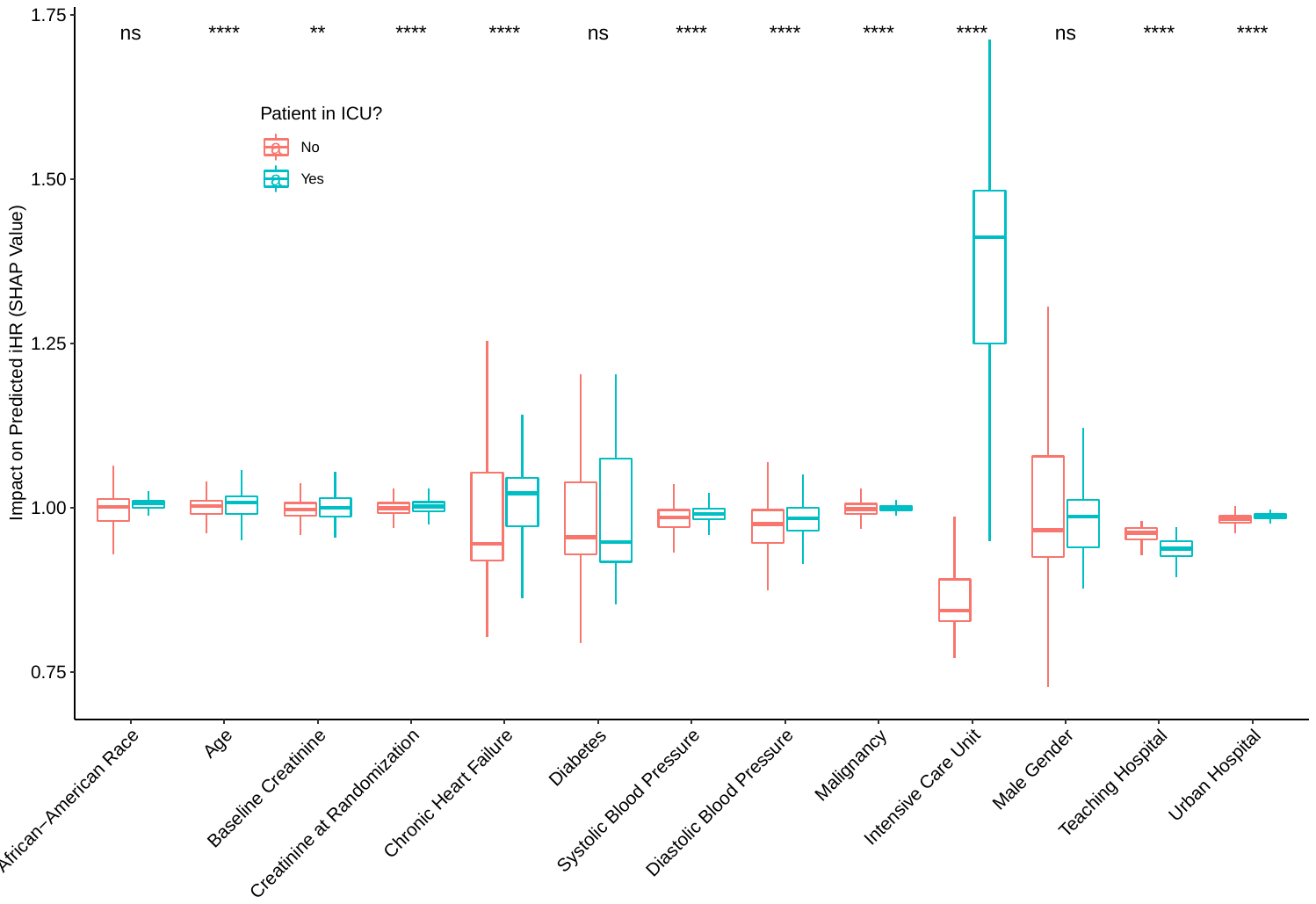

**Supplementary Figure 5.** Interaction between ICU status and the impact of patient characteristics on the treatment effect of alerts (individualized hazard ratio: iHR). These results were from the external validation - the model was trained on the ELAIA-1 trial and feature importance (SHAP values) were calculated using patients from the UPenn trial. NS: not significant (p>0.05); *: p ≤0.05; **: p≤0.01; ***: p≤0.001; ****: p≤0.0001.

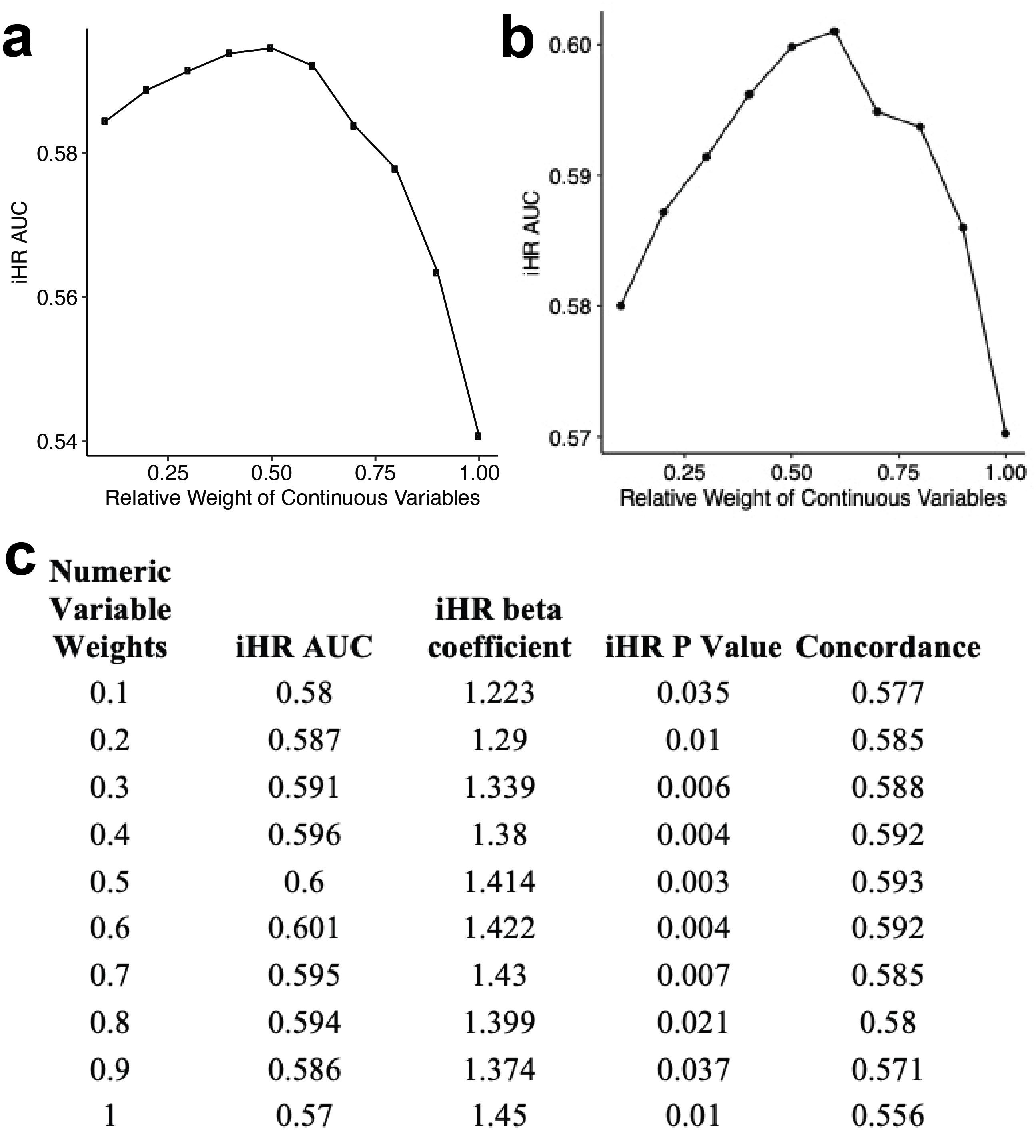

**Supplementary Figure 6.** Hyperparameter tuning of the relative weight of continuous variables for the entire training cohort (a) and the alert group only (b). Additional tuning performance metrics are shown for the alert group (c). The weight of categorial variables corresponded to one minus the weight of continuous variables iHR: individualized hazard ratio; AUC: area under the curve. iHR beta coefficient corresponds to the beta coefficient for the following Cox model: coxph(Surv(time, death) ~ alert*iHR, data=data).

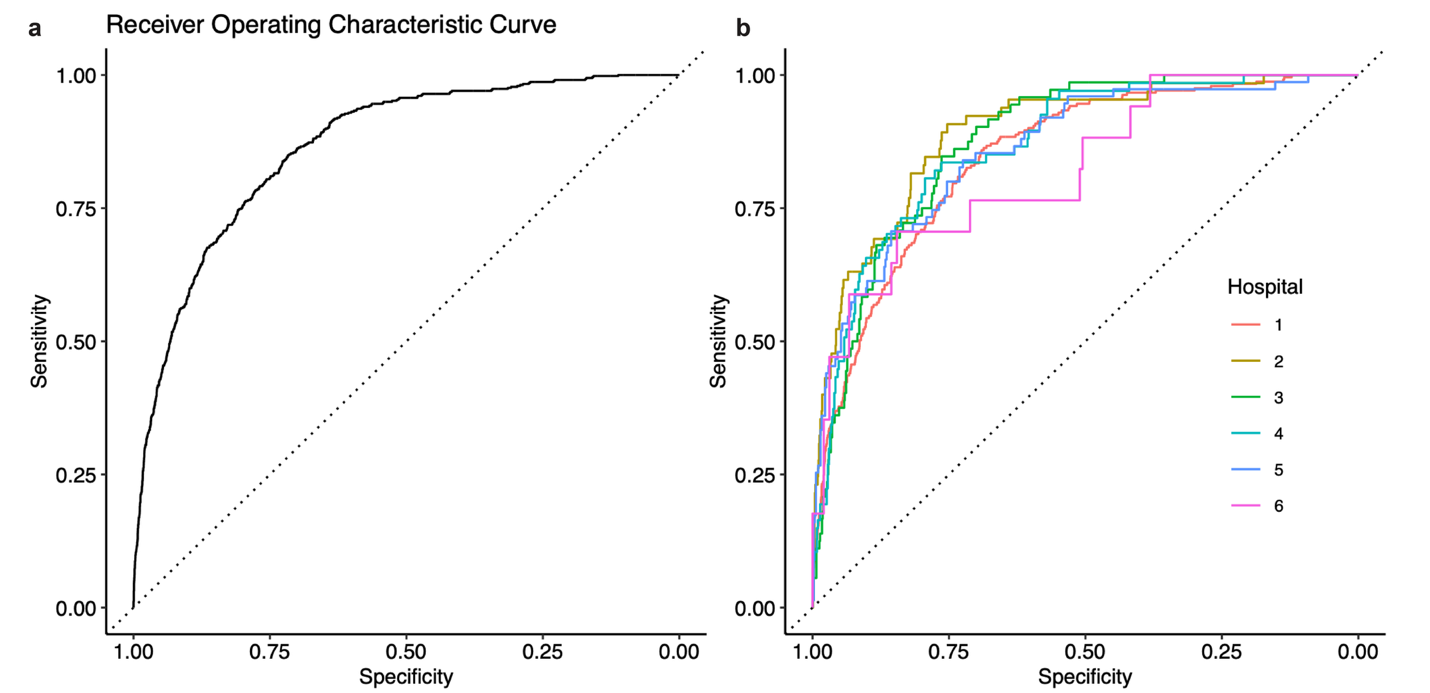

**Supplementary Figure 7.** Receiver operating characteristic curve of the neural network model ensemble. The area under the curve was 0.87 (95% CI: 0.85 to 0.88). The dotted line indicates a random classifier. Model performance at each individual hospital is shown on the right. The best parameters selected to train the NN model were: number of boosting iterations=29, eta=0.1727, gamma=0, max depth=10, minimum child weight=9, and subsample=1.

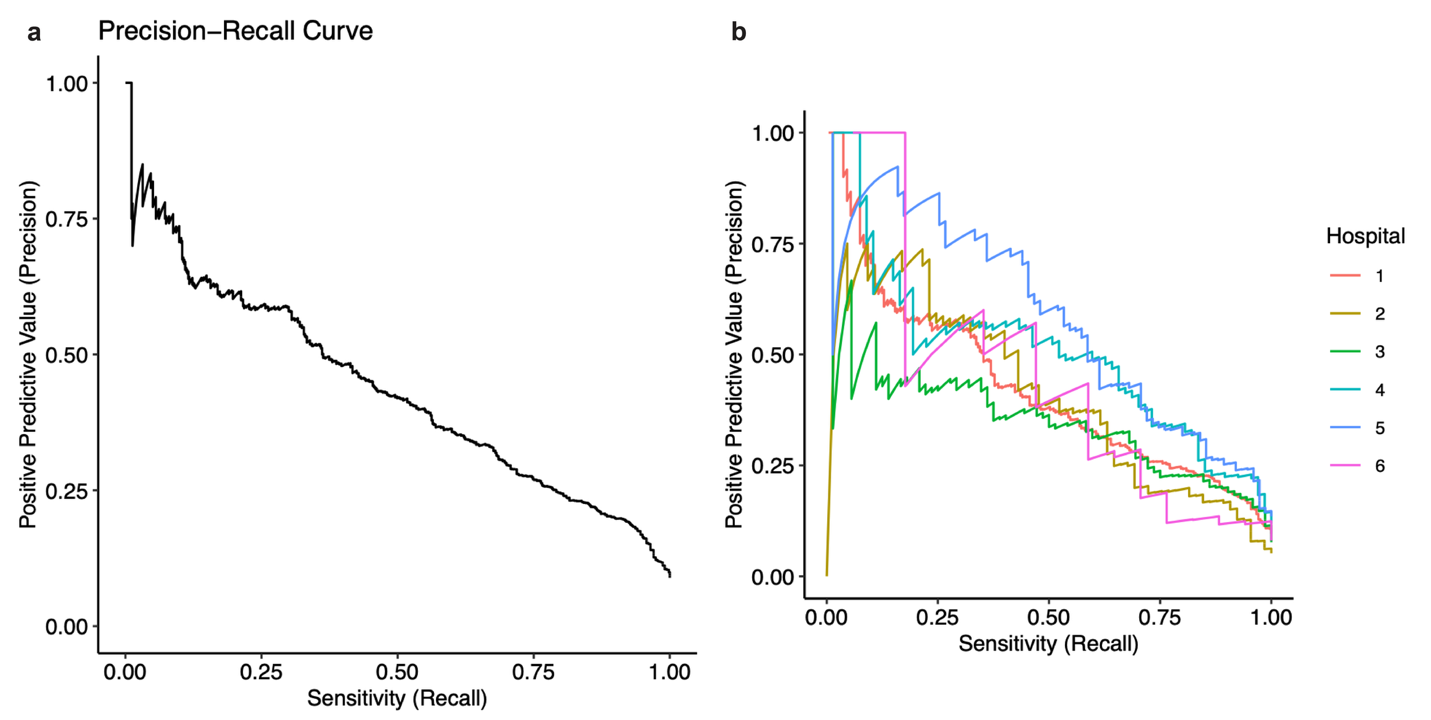

**Supplementary Figure 8.** Precision-recall curve of the deep neural network model ensemble. A) Data across all hospitals. B) Data stratified across individual hospitals. This NN performance indicated the model captured substantial signal of the likelihood a patient would die within 14 days of randomization.

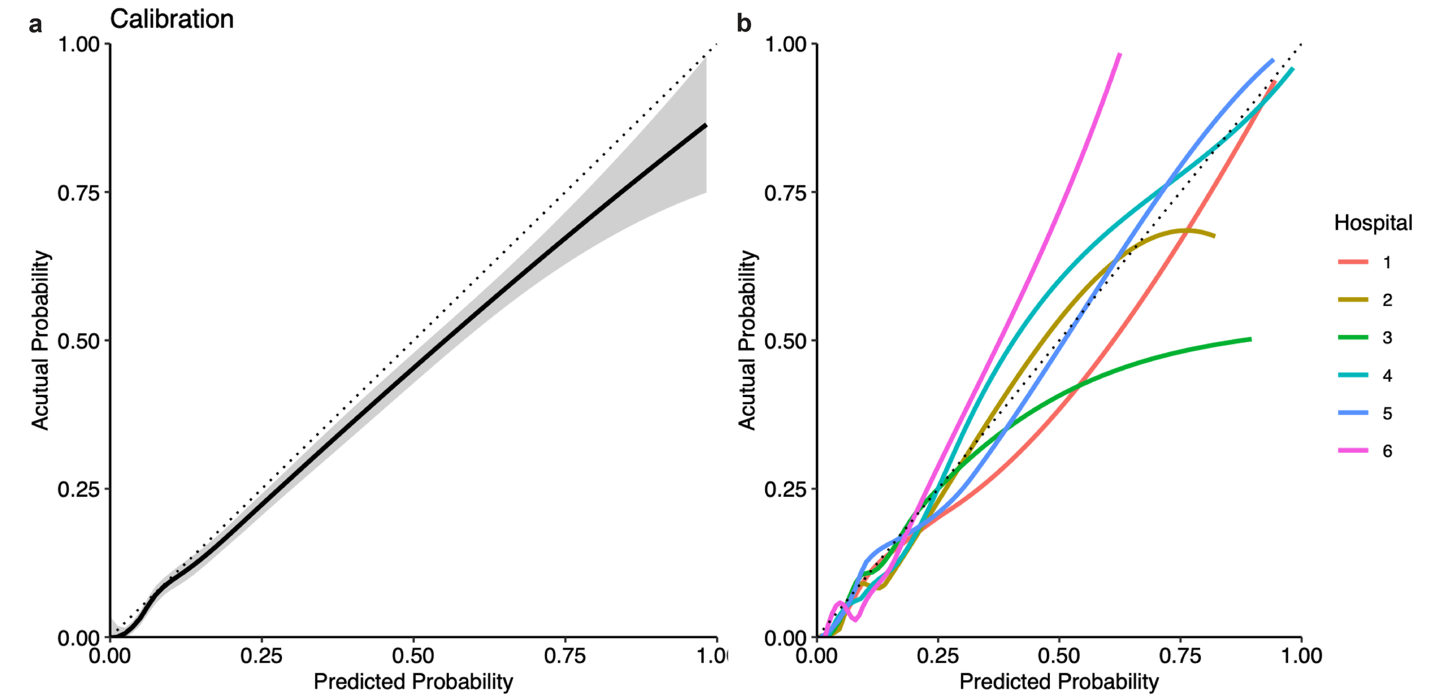

**Supplementary Figure 9.** Non-parametric calibration curve of the neural network’s predicted risk of death within 14 days of randomization. The dotted line indicates perfect calibration. Calibration curves were calculated using local polynomial regression fitting (“loess” option in the “geom_smooth” function in the ggplot2 package in R). This provided evidence near-Level 3 calibration, according the Van Calster et al.,^1^ although there were some deviations within hospitals (shown on the right side of the figure).

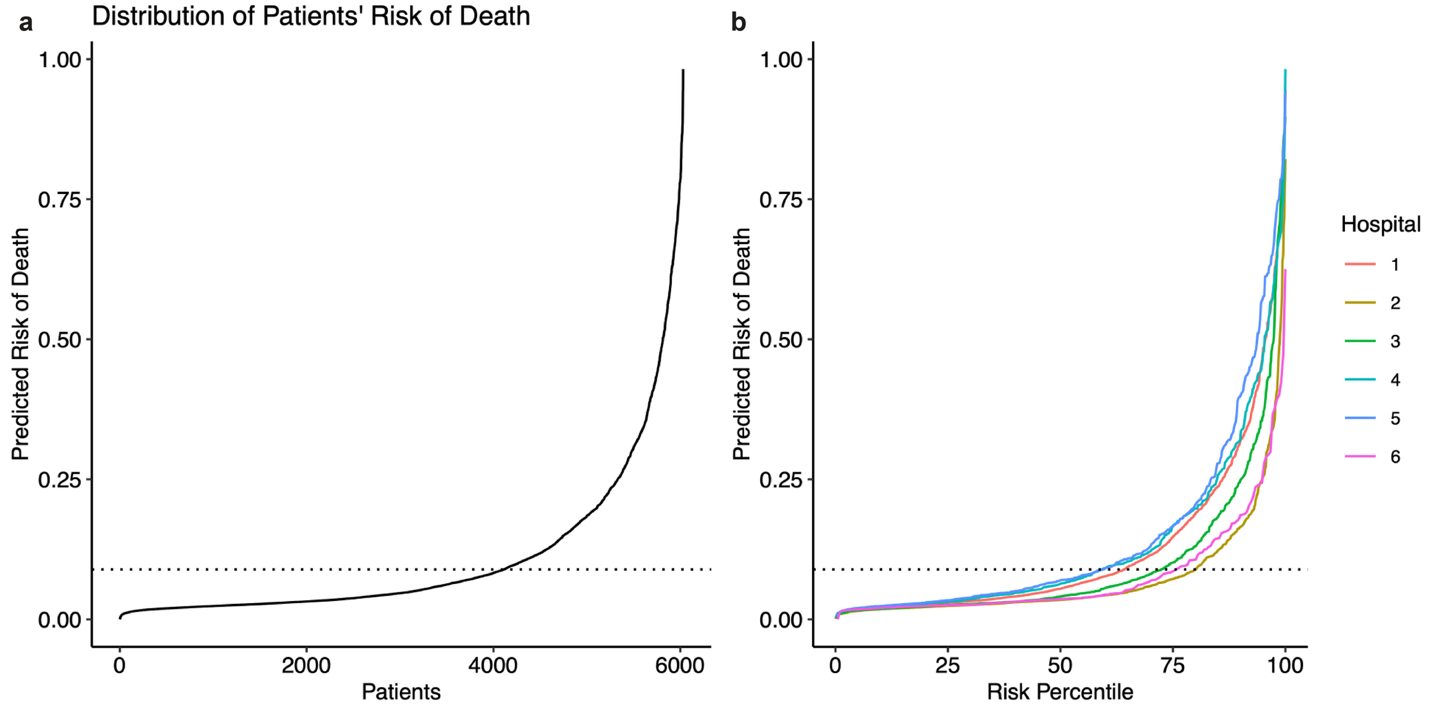

**Supplementary Figure 10.** Distribution of the neural network’s predicted risk of death within 14 days of randomization. The dotted line indicates the observed risk of death for the study population (8.9%). Patients’ predicted risk of mortality was calibrated to the observed risk of mortality and ranged from 0.00 to 0.98 (median: 0.05 [IQR: 0.03 to 0.12]).

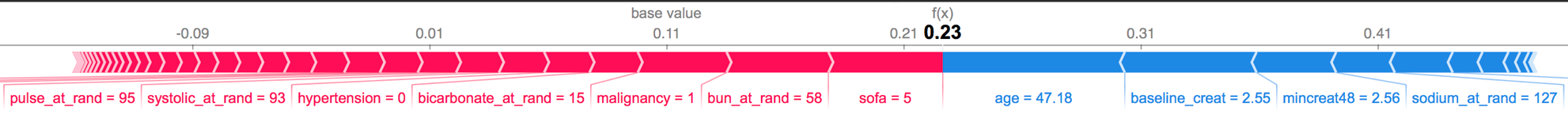

**Supplementary Figure 11.** Force plot showing which features contributed to the predicted risk of death. Plot is for one example patient who did not receive an alert. For example, the predicted risk for this patient started at the population average, 0.11, , and increased to 0.23 based on his specific characteristics: modified Sequential Organ Failure Assessment (mSOFA) score = 5, blood urea nitrogen (BUN) = 58 mg/dl, bicarbonate = 15 mmol/l, systolic blood pressure = 95 mmHG, and history of a malignancy. The model considered his age, 47 years old, creatinine = 2.55 mg/dl, and sodium = 127 mEg/l to be factors that lowered his predicted risk of death.^2^ This patient was randomized to the usual care group and, in this case, not receiving an alert added 0.01 to his predicted risk of mortality.

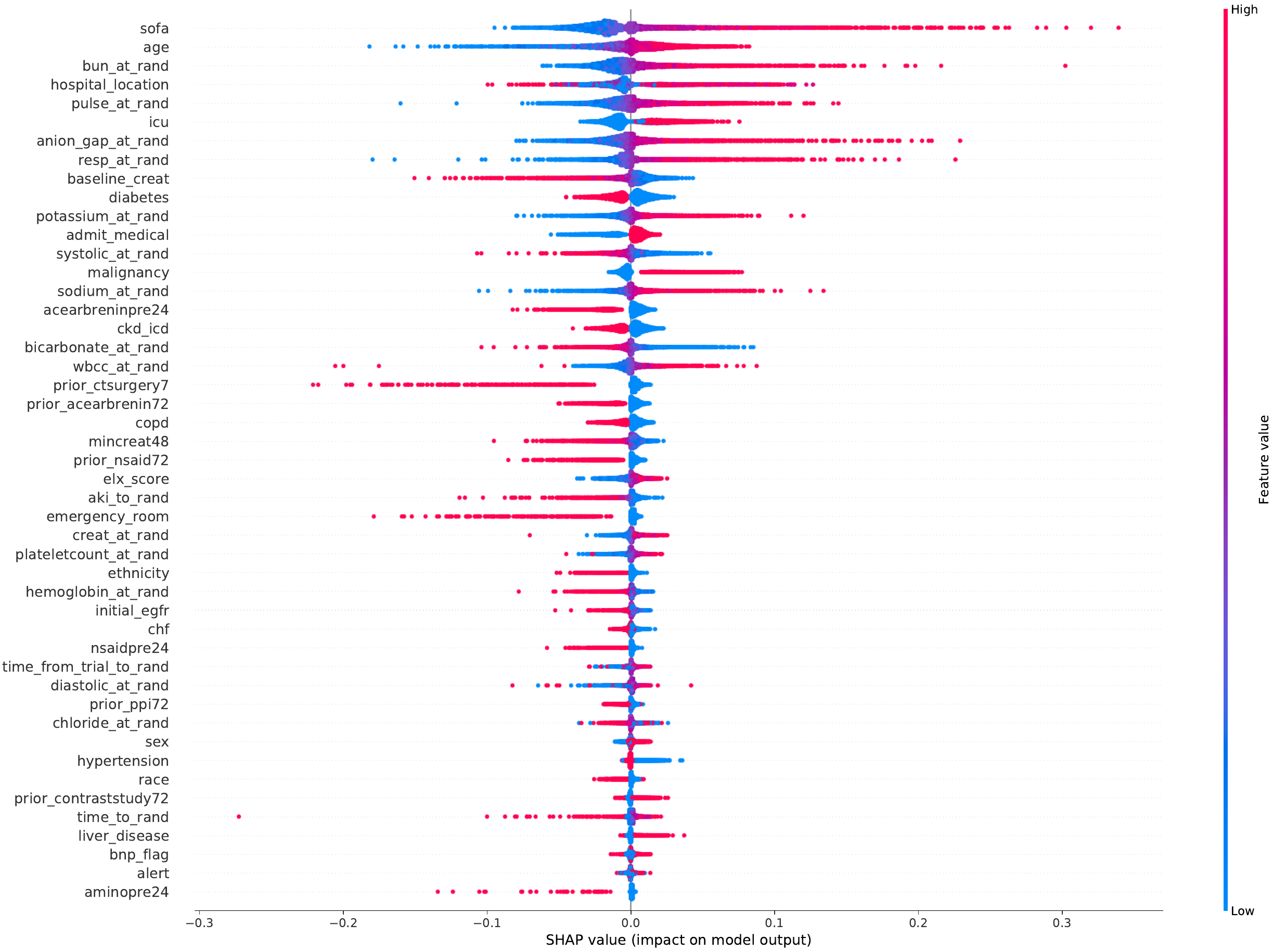

**Supplementary Figure 12.** Summary of feature impact on patients’ predicted risk of death. Each dot represents one patient. Patients that had a low feature value were colored in blue, while high feature values were colored in red. SHAP values (x-axis) correspond to the impact that features had on the predicted risk of death. Variables at the top had the largest impact, while variables at the bottom had the lowest impact on patients’ risk of death.

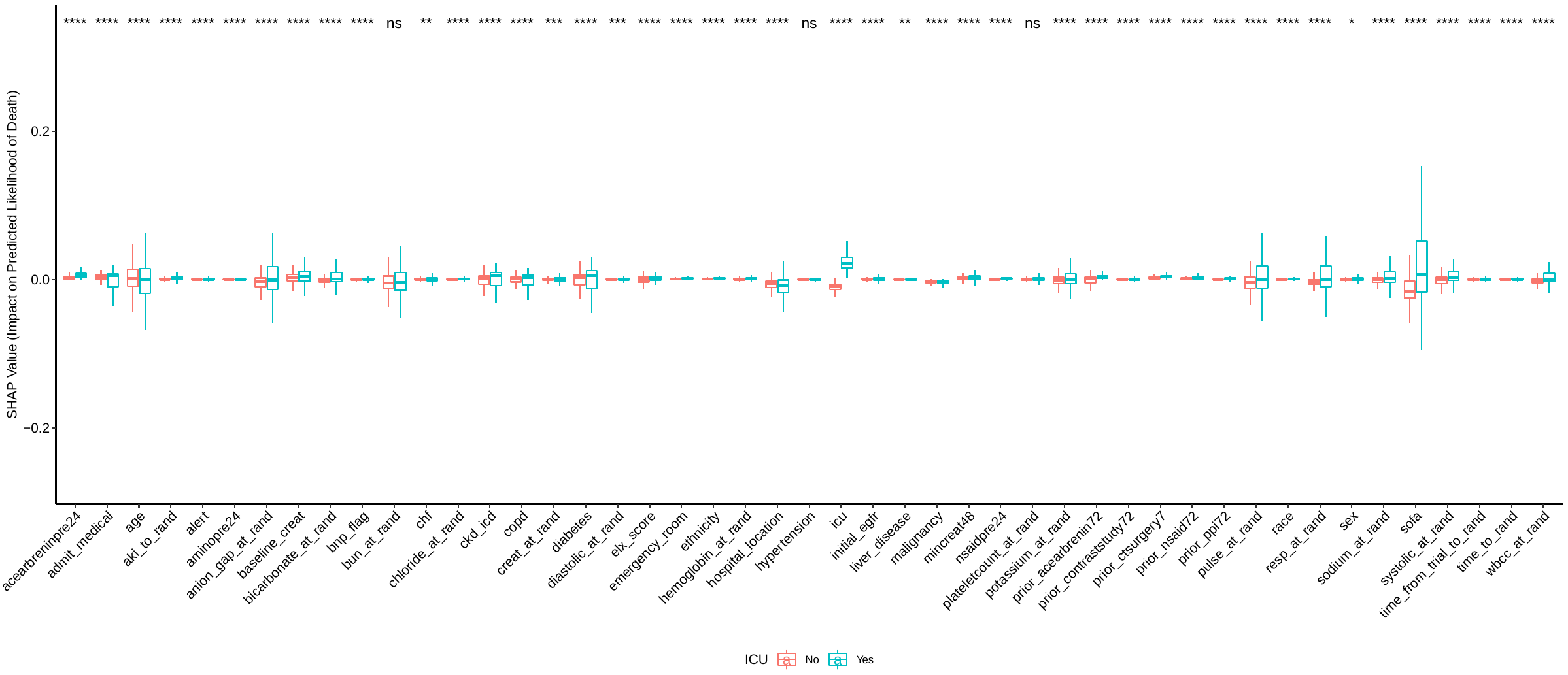

**Supplementary Figure 13.** Interaction between patient location in the intensive care unit (ICU) and the impact of patient characteristics on the predicted risk of mortality. Comparisons were performed using two-sided Wilcoxon rank sum tests. NS: not significant (p>0.05); *: p ≤0.05; **: p≤0.01; ***: p≤0.001; ****: p≤0.0001.

**Supplementary Note 1:**

MEDLINE MESH: ("Acute Kidney Injury"[MeSH] OR "acute kidney injury"[tiab] OR AKI[tiab]) AND (alert[tiab] OR alerts[tiab] OR e-alert[tiab] OR "electronic alert"[tiab] OR "automated alert"[tiab] OR "clinical decision support"[tiab] OR "electronic health record"[tiab]) AND (randomized[tiab] OR randomised[tiab] OR "randomized controlled trial"[pt])

**Supplementary Note 2: PRISMA-IPD Checklist of items to include when reporting a systematic review and meta-analysis of individual participant data (IPD)**

| **PRISMA-IPD**  **Section/topic** | **Item No** | **Checklist item** | **Reported on page** |
| --- | --- | --- | --- |
| **Title** | | | |
| Title | 1 | Identify the report as a systematic review and meta-analysis of individual participant data. | X |
| **Abstract** | | | |
| Structured summary | 2 | Provide a structured summary including as applicable: | 2 |
|  |  | **Background**: state research question and main objectives, with information on participants, interventions, comparators and outcomes. |  |
|  |  | **Methods**: report eligibility criteria; data sources including dates of last bibliographic search or elicitation, noting that IPD were sought; methods of assessing risk of bias. |  |
|  |  | **Results**: provide number and type of studies and participants identified and number (%) obtained; summary effect estimates for main outcomes (benefits and harms) with confidence intervals and measures of statistical heterogeneity. Describe the direction and size of summary effects in terms meaningful to those who would put findings into practice. |  |
|  |  | **Discussion:** state main strengths and limitations of the evidence, general interpretation of the results and any important implications. |  |
|  |  | **Other:** report primary funding source, registration number and registry name for the systematic review and IPD meta-analysis. |  |
| **Introduction** | | | |
| Rationale | 3 | Describe the rationale for the review in the context of what is already known. | 3 |
| Objectives | 4 | Provide an explicit statement of the questions being addressed with reference, as applicable, to participants, interventions, comparisons, outcomes and study design (PICOS). Include any hypotheses that relate to particular types of participant-level subgroups. | 3-4 |
| **Methods** | | | |
| Protocol and registration | 5 | Indicate if a protocol exists and where it can be accessed. If available, provide registration information including registration number and registry name. Provide publication details, if applicable. | 19 |
| Eligibility criteria | 6 | Specify inclusion and exclusion criteria including those relating to participants, interventions, comparisons, outcomes, study design and characteristics (e.g. years when conducted, required minimum follow-up). Note whether these were applied at the study or individual level i.e. whether eligible participants were included (and ineligible participants excluded) from a study that included a wider population than specified by the review inclusion criteria. The rationale for criteria should be stated. | 19 |
| Identifying studies - information sources | 7 | Describe all methods of identifying published and unpublished studies including, as applicable: which bibliographic databases were searched with dates of coverage; details of any hand searching including of conference proceedings; use of study registers and agency or company databases; contact with the original research team and experts in the field; open adverts and surveys. Give the date of last search or elicitation. | 10 |
| Identifying studies - search | 8 | Present the full electronic search strategy for at least one database, including any limits used, such that it could be repeated. | Supplementary Note 1 |
| Study selection processes | 9 | State the process for determining which studies were eligible for inclusion. | 19 |
| Data collection processes | 10 | Describe how IPD were requested, collected and managed, including any processes for querying and confirming data with investigators. If IPD were not sought from any eligible study, the reason for this should be stated (for each such study). | 12-19 |
|  |  | If applicable, describe how any studies for which IPD were not available were dealt with. This should include whether, how and what aggregate data were sought or extracted from study reports and publications (such as extracting data independently in duplicate) and any processes for obtaining and confirming these data with investigators. |  |
| Data items | 11 | Describe how the information and variables to be collected were chosen. List and define all study level and participant level data that were sought, including baseline and follow-up information. If applicable, describe methods of standardizing or translating variables within the IPD datasets to ensure common scales or measurements across studies. | 19 |
| IPD integrity | A1 | Describe what aspects of IPD were subject to data checking (such as sequence generation, data consistency and completeness, baseline imbalance) and how this was done. | 19 |
| Risk of bias assessment in individual studies. | 12 | Describe methods used to assess risk of bias in the individual studies and whether this was applied separately for each outcome. If applicable, describe how findings of IPD checking were used to inform the assessment. Report if and how risk of bias assessment was used in any data synthesis. | 19 |
| Specification of outcomes and effect measures | 13 | State all treatment comparisons of interests. State all outcomes addressed and define them in detail. State whether they were pre-specified for the review and, if applicable, whether they were primary/main or secondary/additional outcomes. Give the principal measures of effect (such as risk ratio, hazard ratio, difference in means) used for each outcome. | 19-22 |
| Synthesis methods | 14 | Describe the meta-analysis methods used to synthesise IPD. Specify any statistical methods and models used. Issues should include (but are not restricted to):   - Use of a one-stage or two-stage approach. - How effect estimates were generated separately within each study and combined across studies (where applicable). - Specification of one-stage models (where applicable) including how clustering of patients within studies was accounted for. - Use of fixed or random effects models and any other model assumptions, such as proportional hazards. - How (summary) survival curves were generated (where applicable). - Methods for quantifying statistical heterogeneity (such as I^2^ and t^2^). - How studies providing IPD and not providing IPD were analysed together (where applicable). - How missing data within the IPD were dealt with (where applicable). | 19-22 |
| Exploration of variation in effects | A2 | If applicable, describe any methods used to explore variation in effects by study or participant level characteristics (such as estimation of interactions between effect and covariates). State all participant-level characteristics that were analysed as potential effect modifiers, and whether these were pre-specified. | 19-22 |
| Risk of bias across studies | 15 | Specify any assessment of risk of bias relating to the accumulated body of evidence, including any pertaining to not obtaining IPD for particular studies, outcomes or other variables. | 19-22 |
| Additional analyses | 16 | Describe methods of any additional analyses, including sensitivity analyses. State which of these were pre-specified. | 12-22 |
| **Results** | | | |
| Study selection and IPD obtained | 17 | Give numbers of studies screened, assessed for eligibility, and included in the systematic review with reasons for exclusions at each stage. Indicate the number of studies and participants for which IPD were sought and for which IPD were obtained. For those studies where IPD were not available, give the numbers of studies and participants for which aggregate data were available. Report reasons for non-availability of IPD. Include a flow diagram. | 4 |
| Study characteristics | 18 | For each study, present information on key study and participant characteristics (such as description of interventions, numbers of participants, demographic data, unavailability of outcomes, funding source, and if applicable duration of follow-up). Provide (main) citations for each study. Where applicable, also report similar study characteristics for any studies not providing IPD. | 4-9 |
| IPD integrity | A3 | Report any important issues identified in checking IPD or state that there were none. | 4 |
| Risk of bias within studies | 19 | Present data on risk of bias assessments. If applicable, describe whether data checking led to the up-weighting or down-weighting of these assessments. Consider how any potential bias impacts on the robustness of meta-analysis conclusions. | 4 |
| Results of individual studies | 20 | For each comparison and for each main outcome (benefit or harm), for each individual study report the number of eligible participants for which data were obtained and show simple summary data for each intervention group (including, where applicable, the number of events), effect estimates and confidence intervals. These may be tabulated or included on a forest plot. | 4-9 |
| Results of syntheses | 21 | Present summary effects for each meta-analysis undertaken, including confidence intervals and measures of statistical heterogeneity. State whether the analysis was pre-specified, and report the numbers of studies and participants and, where applicable, the number of events on which it is based. | 4-9 |
|  |  | When exploring variation in effects due to patient or study characteristics, present summary interaction estimates for each characteristic examined, including confidence intervals and measures of statistical heterogeneity. State whether the analysis was pre-specified. State whether any interaction is consistent across trials. |  |
|  |  | Provide a description of the direction and size of effect in terms meaningful to those who would put findings into practice. |  |
| Risk of bias across studies | 22 | Present results of any assessment of risk of bias relating to the accumulated body of evidence, including any pertaining to the availability and representativeness of available studies, outcomes or other variables. | 4 |
| Additional analyses | 23 | Give results of any additional analyses (e.g. sensitivity analyses). If applicable, this should also include any analyses that incorporate aggregate data for studies that do not have IPD. If applicable, summarise the main meta-analysis results following the inclusion or exclusion of studies for which IPD were not available. | 4-9 |
| **Discussion** | | | |
| Summary of evidence | 24 | Summarise the main findings, including the strength of evidence for each main outcome. | 9-12 |
| Strengths and limitations | 25 | Discuss any important strengths and limitations of the evidence including the benefits of access to IPD and any limitations arising from IPD that were not available. | 9-12 |
| Conclusions | 26 | Provide a general interpretation of the findings in the context of other evidence. | 12 |
| Implications | A4 | Consider relevance to key groups (such as policy makers, service providers and service users). Consider implications for future research. | 12 |
| **Funding** | | | |
| Funding | 27 | Describe sources of funding and other support (such as supply of IPD), and the role in the systematic review of those providing such support. | 24 |

**A1 – A3 denote new items that are additional to standard PRISMA items. A4 has been created as a result of re-arranging content of the standard PRISMA statement to suit the way that systematic review IPD meta-analyses are reported.**

© Reproduced with permission of the PRISMA IPD Group, which encourages sharing and reuse for non-commercial purposes

**SUPPLEMENTARY NOTE 3**

Weighting continuous and categorical variables 50:50 optimized discrimination for the individualized hazard ratios, as determined by the area under the receiver operating characteristic curve for the ability to predict death within 14 days (Supplementary Figure 6). This meant that each of the five continuous variables contributed 10% when calculating Gower’s distance (50% total weight for continuous variables) while each of the eight categorical variables contributed 6.25% (50% total weight for categorical variables).

Using the full set of baseline features available in ELAIA-1, the NN predicted death with an AUC of 0.87 (95% CI: 0.85 to 0.88; Supplementary Figures 7 and 8). This performance indicated the model captured substantial signal of the likelihood a patient would die within 14 days of randomization. Patients’ predicted risk of mortality was calibrated to the observed risk of mortality (Supplementary Figure 9) and ranged from 0.00 to 0.98 (median: 0.05 [IQR: 0.03 to 0.12]; Supplementary Figure 10). Predictions for the alert and usual-care groups were balanced at the population level (difference in means: 0.00 [95% CI: -0.007 to 0.007]; p=0.45), suggesting that the randomization procedure effectively balanced patient covariates across the intervention groups. Using the limited set of baseline features (patient sex, race, age, blood pressure, baseline creatinine, creatinine at the time of randomization, congestive heart failure (CHF) diagnosis, diabetes diagnosis, any malignancy, hospitalization in the intensive care unit, and hospital type), AUC for predicting baseline risk of death was 0.764 (95% CI: 0.745 to 0.784). Our ensemble of 100 NNs performed better than a single NN with the same hyperparameters (AUC = 0.757 [95% CI: 0.737 to 0.777]; p = 0.02).

Knowledge encoded in the NNs was interrogated using SHAP (Shapley Additive exPlanations) values. SHAP values indicated the potential impact a specific feature value had on an individual patient’s risk of death. For example, the predicted risk of mortality for the patient shown in Supplementary Figure 11 started at the population average, 0.11, and increased to 0.23 based on his specific characteristics: modified Sequential Organ Failure Assessment (mSOFA) score = 5, blood urea nitrogen (BUN) = 58 mg/dl, bicarbonate = 15 mmol/l, systolic blood pressure = 95 mmHG, and history of a malignancy. The model considered his age, 47 years old, creatinine = 2.55 mg/dl, and sodium = 127 mEg/l to be factors that lowered his predicted risk of death.^2^ This patient was randomized to the usual care group and, in this case, not receiving an alert added 0.01 to his predicted risk of mortality.

A global summary of how features impacted the NN’s predicted risk of death is shown in Supplementary Figure 12.^3^ mSOFA scores, age, BUN, anion gap, creatinine, potassium, pulse, and systolic blood pressure had the largest impact on the model’s estimated risk of death. Patient location in the intensive care unit modulated the effect nearly every feature’s impact on the predicted risk of death (Supplementary Figure 13).
